# Effect of a community-based behavioural intervention bundle to improve antibiotic use and patient management in Burkina Faso and DR Congo: a cluster randomised controlled trial

**DOI:** 10.64898/2025.12.15.25342146

**Authors:** Brecht Ingelbeen, Daniel Valia, Bijou Mbangi, Esther van Kleef, Linda Campbell, Stéphane Juste Kouanda, César-Augustin Mambuku Khoso Muaka, Eric Wendpouiré Tiendrébeogo, Aminata Welgo, Veerle Bertels, Steven Declercq, Bram Riems, Marie Meudec, Edwin Wouters, Ben S Cooper, Delphin Mavinga Phanzu, Halidou Tinto, Marianne A.B. van der Sande, CABU-EICO study group

## Abstract

**Background:** Increasing Watch-group antibiotic use may be contributing to antimicrobial resistance burden in sub-Saharan Africa. We evaluated the effect of a community-based, co-created intervention bundle targeting all community-level healthcare providers and communities they serve, on Watch-group antibiotic use and patient management.

**Methods:** In a cluster-randomised, controlled trial in Burkina Faso (BF) and Democratic Republic of Congo (DRC), 44 villages with ≥500 inhabitants and ≥1 health centre or medicine vendor were randomly allocated 1:1 to intervention or control arms, using the RAND function in Excel. Over nine months, three intervention rounds consisted of community health education campaigns and educational/feedback sessions with providers, introducing AWaRe Book guidance for infections with highest antibiotic use. We measured baseline-to-post-intervention changes in Watch-group antibiotic use through repeated patient surveys (100 per provider per village), cluster-adjusted and offset for healthcare utilisation (primary outcome), and patient management through simulated patient visits (secondary outcome). Field workers conducting patient surveys and simulated patient visits were masked. CABU-EICO was registered on clinicaltrials.gov/study/NCT05378880.

**Findings:** At baseline (Oct 26, 2022 to Mar 13, 2023), 5532 patients were surveyed (3558 in BF; 1974 in DRC); post-intervention (Nov 6, 2023 to Apr 3, 2024), 4898 patients (3180 in BF; 1718 in DRC). Surveys were completed at 63 health centres, 60 pharmacies, and 41 informal vendors. A total of 1092 simulated patient visits were completed across both periods. Weighted prevalence of Watch-group antibiotic use decreased from 26·8% (95%CI 8·8-44·8) to 17·1% (95%CI 7·7-26·5) in intervention and increased from 13·4% (95%CI4·8-22) to 21.2% (95%CI8·9-34) in control clusters; adjusted prevalence ratio 0·33 (95%CI 0·14-0·78). Changes in patient management scores were limited.

**Interpretation:** The behavioural intervention bundle substantially reduced Watch-group antibiotic use and did not negatively impact patient management, highlighting the potential of antibiotic use improvements across healthcare providers. Reduced community-level use of broad-spectrum antibiotics could help slow community-acquired pathogens’ increasing resistance to clinically important antibiotics.

**Funding:** JPI-AMR, Research Foundation-Flanders

**Research in context:** *Evidence before this study:* Evidence on the effectiveness of interventions to improve antibiotic use in primary care in low- and middle-income countries (LMIC) is heterogenous, in terms of intervention components and effect sizes. A systematic review of behavioural interventions in LMIC between 2001 and 2019 analysed 13 studies in health centres, private clinics, and pharmacies. We complemented the review with a search on Pubmed for interventional studies in primary care or community-level in LMIC since 2019, using (“Anti-Bacterial Agents” OR “Antimicrobial Stewardship”) AND (“Guideline Adherence” OR “Education, Medical, Continuing” OR “Feedback” OR “Decision Support Systems, Clinical” OR “Drug Utilization Review” OR “Clinical Protocols” OR “prescription audit”) AND (“Ambulatory Care” OR “Primary Health Care” OR “Outpatients” OR “Pharmacies” OR “Nonprescription Drugs”). Most interventions consisted of educational sessions on treatment guidance, with or without feedback, or combined with clinical algorithms. Studies conducted in Vietnam, Sudan, Tanzania, India, Kenya, and China reported reductions in inappropriate antibiotic prescribing ranging by 16 (audit/feedback alone, Sudan) to 55 percentage point (84.3% to 15.4%, Tanzania), with the largest effects consistently seen in bundle interventions combining education, audit and feedback, and regulatory enforcement. Evidence concerning interventions among informal healthcare providers is scarcer: provision of educational sessions for informal medicine vendors in India was not linked to a change in prevalence of antibiotic use but did improve patient management. A systematic review of studies in sub-Saharan Africa published up to 2020 found that populations frequently self-medicated with antibiotics, obtained without prescription from community pharmacies or (informal) medicine vendors.

*Added value of this study:* The study evaluated a co-created intervention bundle that simultaneously targeted both antibiotic dispensing by healthcare providers or vendors and the demand from the communities they serve, which, to our knowledge, no previous published study did. Three intervention rounds consisted of health education campaigns with communities and educational and feedback sessions with providers, introducing AWaRe Book guidance for infections with highest antibiotic use. By including all community-level providers and vendors in both the intervention and its evaluation, and by adjusting community-level antibiotic-use prevalence for healthcare utilisation, we were able to estimate effects on community-wide antibiotic use. The intervention study was conducted in two sites in Sub-Saharan Africa, in Burkina Faso and DR Congo, with differences in antibiotic dispensing and in available healthcare providers. That the relative effect size was comparable between sites, despite baseline differences, supports external validity of the intervention bundle’s effectiveness. We used simulated patient visits for five priority infections to assess changes in patient management and antibiotic dispensing. This approach enabled direct comparison of history taking, examination practices, and dispensing across provider types and intervention arms, offering insight into how the intervention influenced antibiotic use and clinical management. Together with detailed sub-analyses by infection and provider type, and qualitative interviews with providers and community members, the evaluation identified intervention components which appeared to be the most effective and could be prioritised in future, more targeted interventions. Finally, this is the first experimental study to evaluate an intervention informed by the WHO AWaRe Antibiotic Book, which was published in December 2022, only two months before the study started..

*Implications of all the available evidence:* Contextualised behavioural interventions based on existing treatment guidance, focusing on a limited number of common primary-care infections, can substantially reduce antibiotic use and improve the selection of appropriate antibiotics, potentially slowing the increasing resistance of community-acquired pathogens to clinically important antibiotics. Reductions in antibiotic use were greatest in clinics and health centres, with private clinics contributing importantly to the reduction. Reducing antibiotic sales from medicine vendors proved difficult without stronger regulatory measures.

## Introduction

Mortality attributable to bacterial antimicrobial resistance (AMR) is estimated to be highest in sub-Saharan Africa.^1,2^ Causative pathogens are predominantly community-acquired, such as *Streptococcus pneumoniae, Escherichia coli*, and (non-)typhoid *Salmonella*, and frequently exhibit resistance to Access and Watch-group antibiotics that are essential to treat most bacterial infections.^1,3,4^ High resistance levels may stem from both community- and healthcare-level drivers such as inappropriate antibiotic use leading to selective pressure favouring resistant bacteria, and poor sanitation leading to high rates of transmission between hosts and environmental reservoirs.^5–7^ Core objectives of the Global Action Plan on AMR are, accordingly, to optimize the use of antimicrobials and to reduce the incidence of infection through effective sanitation, hygiene and infection prevention measures.^8^

In 2022, the World Health Organization (WHO) introduced the AWaRe Antibiotic Book, evidence-based treatment guidelines for empiric prescribing in primary care and hospitals.^9^ Its guidance for 20 common infections in primary care promotes symptomatic care without antibiotics and the use of Access over Watch-group antibiotics. Watch-group antibiotics are more likely to select for high-priority multidrug resistant organisms.^10^ For most minor infections (mainly upper respiratory tract infections), no antibiotic use is recommended. Watch-group antibiotics are only recommended for acute severe bloody diarrhoea, enteric fever, and certain sexually transmitted infections. The UN General Assembly committed in September 2024 to expand by 2030 the use of Access antibiotics to at least 70% of overall human antibiotic use, by strengthening stewardship programmes.^11^

Unnecessary or suboptimal antibiotic use can arise from limited awareness of antimicrobial resistance, limited diagnostic capacity, and frequent self-medication in low- and middle-income countries (LMIC).^12,13^ In sub-Saharan Africa, antibiotic use per capita is increasing, but overall it is not thought to be higher than in most industrialised countries.^14–16^ Nevertheless, previous patient surveys in rural Burkina Faso and DR Congo indicated that most current use of antibiotics in primary care is for conditions for which (Watch-group) antibiotics are not recommended: respectively 69% and 75% of Watch-group-antibiotic use could be replaced by Access antibiotics or no antibiotic use, if treatment would have been according to guidance from the AWaRe Antibiotic Book.^15,17^

Evidence about effectiveness of interventions to improve community-level or primary care antibiotic use in LMIC is limited and heterogeneous, in terms of intervention components, healthcare providers targeted, and effect size. Intervention bundles combining enabling interventions (e.g., education, guidelines) with persuasive interventions (e.g., dispensing audits and feedback) appeared more effective than standalone interventions.^18,19^

The CABU-EICO project developed, implemented and evaluated a community-based intervention bundle targeting all medicine providers (supply) and communities they serve (demand), to improve human antibiotic use and reduce transmission through optimised antibiotic use and hygiene improvements in communities in two health districts in Burkina Faso and DR Congo.^20^ We evaluated the intervention’s effect on Watch-group and overall antibiotic use and on patient management across providers.

## Methods

### Study design

We conducted a cluster-randomised, controlled trial in 44 villages and neighbourhoods in two health districts in two countries: Kimpese in DR Congo and Nanoro in Burkina Faso. Study protocols were approved by Université Protestante au Congo Ethics Committee (CEUPC0098), Burkina Faso Ethics Committee for Health Research (2022-03-050), and Antwerp University Hospital Ethics Committee (3456, 3363). Medicine providers participating to the intervention or evaluation surveys provided written informed consent. Village leaders provided oral consent for villages’ participation to the health education campaign. Patients ≥18 years participating to patient surveys and adult household members participating in household surveys provided written informed consent; for patients<18 years, parents or caretakers provided written informed consent, with assent of patients 14-17 years old. The trial was registered 13 May 2022 on clinicaltrials.gov/study/NCT05378880.

### Clusters

Village or neighbourhood was the unit of randomisation because the intervention was targeting all community-level healthcare providers or medicine dispensers and surrounding communities. In Kimpese, 14 rural villages and eight peri-urban neighbourhoods were selected (appendix pp2). In Nanoro, 22 rural villages were selected. Eligible village or neighbourhood clusters had at least 500 inhabitants, and at least one community-level or primary care provider functioning as the main medicine dispenser of the population of that village/neighbourhood. For peri-urban neighbourhoods, non-neighbouring areas were selected, to avoid contamination of a potential intervention effect from intervention to control clusters. Clusters selected for the trial were different from villages where the intervention was developed. Village leaders and local community-level healthcare providers were visited to assess eligibility and willingness to participate in the study.

### Randomisation

Village/neighbourhood clusters were randomised 1:1 intervention or control (usual care) in four strata. In Nanoro, strata distinguished villages with a primary health centre (public or private clinic) and those without (only medicine vendors, i.e. community pharmacy, medicine store without qualified pharmacist, or informal medicine vendor). In Kimpese, strata distinguished peri-urban neighbourhoods and rural villages. A random number between 0 and 1 was given to each cluster using the RAND function in Excel by a researcher not involved in intervention implementation. Per stratum, clusters with the 50% higher numbers were allocated as intervention clusters; the half with the lower numbers were therefore allocated as control clusters. Field workers conducting simulated patient visits or patient surveys were masked, though inadvertent unmasking (e.g., via presence of printed intervention materials) cannot be completely ruled out.

### Intervention and participants

The study team co-created a behavioural intervention package based on consultation of community members and community-level healthcare providers. Content and form of the intervention bundle were developed and contextualised for each site, based on two exploratory qualitative studies, one unpublished and one published,^21^ interviewing and observing all healthcare providers (public health centres, private clinics, community pharmacies, informal medicine vendors) and community members in four villages per site. Villages for intervention development shared socio-economic and geographic characteristics with trial villages. The intervention was later piloted in these villages, as a way of presenting results of the preparatory research whilst also gaining valuable feedback on the intervention. A day-to-day intervention manual specified format and content of three rounds of educational and feedback sessions with providers and mass health education activities with community members (Table 1, appendix pp5). Sessions with providers focused on four infections accounting for 73% (1047/1444) of any antibiotic use and 66% (210/318) of Watch-group antibiotics in prior patient surveys.^15,17^ Priority infections were: (1) cough including bronchitis, (2) community-acquired pneumonia, (3) diarrhoea and other gastro-intestinal complaints, and (4) acute fever without other discriminating symptoms. We translated, adapted and contextualized treatment guidance for those infections from the 2022 AWaRe Antibiotic Book, while integrating existing Integrated Management of Childhood Illness guidance in children aged <5 years (appendix pp5).

**Table 1.**
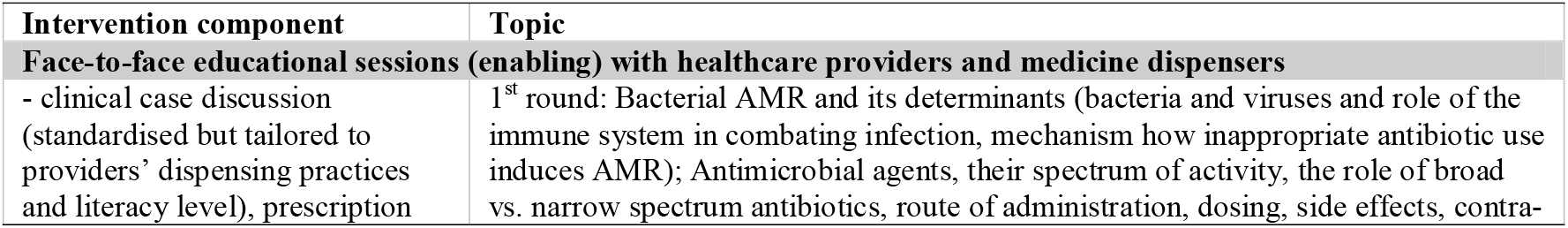
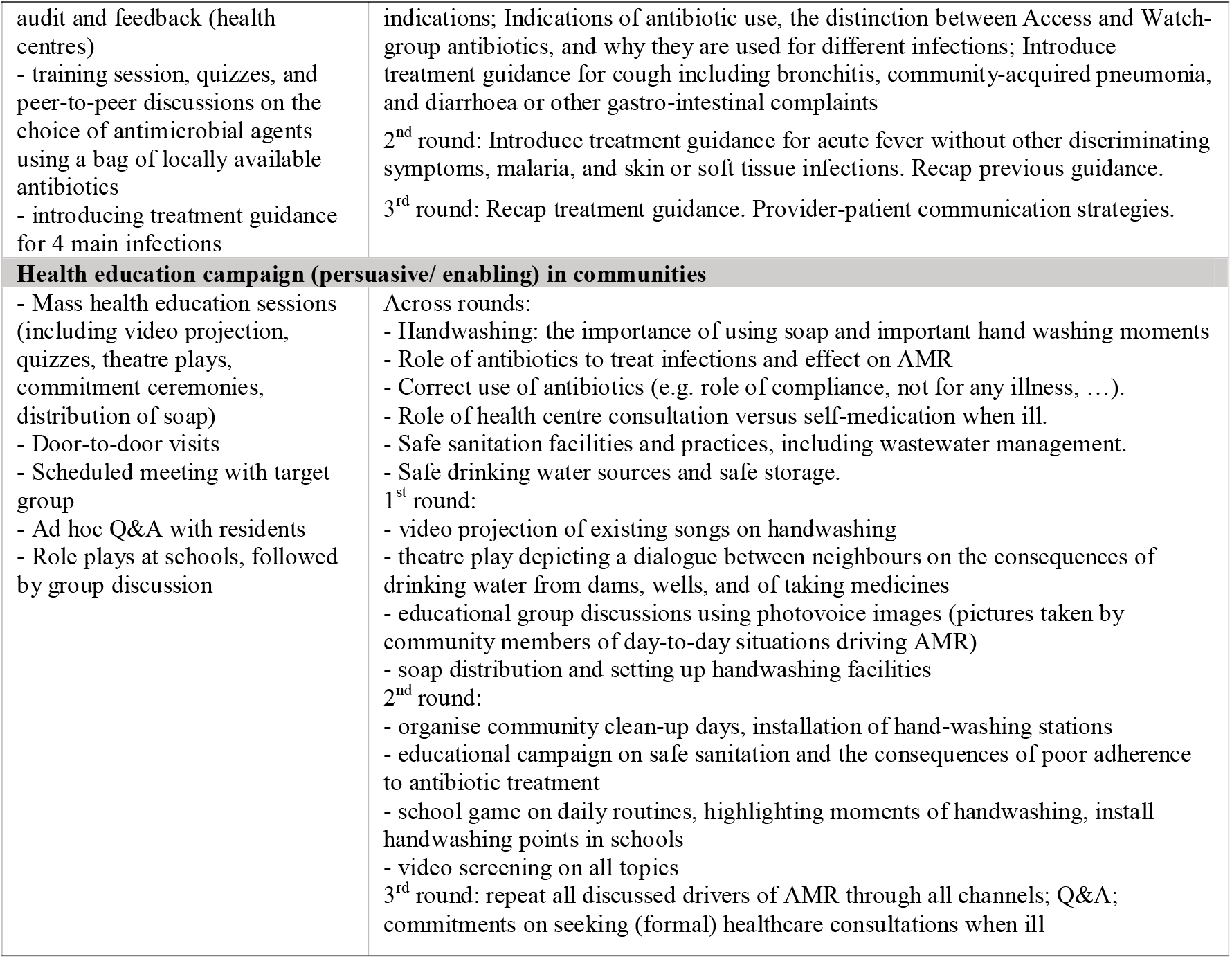
Components and content of the co-created AMR control intervention bundle.

| Intervention component | Topic |
| --- | --- |
| **Face-to-face educational sessions (enabling) with healthcare providers and medicine dispensers** |  |
| - clinical case discussion (standardised but tailored to providers' dispensing practices and literacy level), prescription | 1<sup>st</sup> round: Bacterial AMR and its determinants (bacteria and viruses and role of the immune system in combating infection, mechanism how inappropriate antibiotic use induces AMR); Antimicrobial agents, their spectrum of activity, the role of broad vs. narrow spectrum antibiotics, route of administration, dosing, side effects, contra- |
| audit and feedback (health centres)<br>- training session, quizzes, and peer-to-peer discussions on the choice of antimicrobial agents using a bag of locally available antibiotics<br>- introducing treatment guidance for 4 main infections | indications; Indications of antibiotic use, the distinction between Access and Watch-group antibiotics, and why they are used for different infections; Introduce treatment guidance for cough including bronchitis, community-acquired pneumonia, and diarrhoea or other gastro-intestinal complaints<br>2<sup>nd</sup> round: Introduce treatment guidance for acute fever without other discriminating symptoms, malaria, and skin or soft tissue infections. Recap previous guidance.<br>3<sup>rd</sup> round: Recap treatment guidance. Provider-patient communication strategies. |
| **Health education campaign (persuasive/ enabling) in communities** |  |
| - Mass health education sessions (including video projection, quizzes, theatre plays, commitment ceremonies, distribution of soap)<br>- Door-to-door visits<br>- Scheduled meeting with target group<br>- Ad hoc Q&A with residents<br>- Role plays at schools, followed by group discussion | Across rounds:<br>- Handwashing: the importance of using soap and important hand washing moments<br>- Role of antibiotics to treat infections and effect on AMR<br>- Correct use of antibiotics (e.g. role of compliance, not for any illness, ...).<br>- Role of health centre consultation versus self-medication when ill.<br>- Safe sanitation facilities and practices, including wastewater management.<br>- Safe drinking water sources and safe storage.<br>1<sup>st</sup> round:<br>- video projection of existing songs on handwashing<br>- theatre play depicting a dialogue between neighbours on the consequences of drinking water from dams, wells, and of taking medicines<br>- educational group discussions using photovoice images (pictures taken by community members of day-to-day situations driving AMR)<br>- soap distribution and setting up handwashing facilities<br>2<sup>nd</sup> round:<br>- organise community clean-up days, installation of hand-washing stations<br>- educational campaign on safe sanitation and the consequences of poor adherence to antibiotic treatment<br>- school game on daily routines, highlighting moments of handwashing, install handwashing points in schools<br>- video screening on all topics<br>3<sup>rd</sup> round: repeat all discussed drivers of AMR through all channels; Q&A; commitments on seeking (formal) healthcare consultations when ill |

Three rounds of intervention activities, four days per cluster per round, were implemented by three (Nanoro) to five (Kimpese) educators. One educator with medical background conducted educational and feedback sessions with providers. The intervention and its evaluation were rolled out in a staggered way, visiting – on average – one intervention and one control cluster per week per intervention round, hence completing all within nine months (Jan-Sep 2023 in Nanoro, Apr-Dec 2023 in Kimpese, appendix pp13).

### Outcomes

We report on the primary outcome: the difference in cluster- and provider-weighted prevalence of Watch-group antibiotic use (patients using at least one Watch-group antibiotic) at medicine vendors or health centres, comparing intervention to control clusters, pre- and post-intervention. Another primary outcome, the effect on ESBL-*E. coli* colonisation, based on repeatedly collected stool samples and recorded water access, sanitation, and hygiene practices, is reported elsewhere.^22^ Secondary outcomes were the difference in prevalence of any antibiotic use, the difference in pre-to post-intervention change in patient management score (by simulated infection, per provider), and pathways through which interventions improve the quality of care and treatment outcomes. Exploratory post-hoc outcomes were subgroup analyses by type of provider, by type of infection, the change in per capita antibiotic use, and the change in proportion of non-recommended antibiotic use.

At baseline and post-intervention (12 months later, to account for seasonality), outcomes were recorded in three surveys. First, patient surveys recorded use of systemic antibiotics during or following patient visits to health centres, private clinics, community pharmacies, and informal vendors. Second, household surveys recorded healthcare visits of any household member in the previous month and previous three months, used to estimate the provider-specific rate of healthcare utilisation. Third, simulated patients visits were conducted to assess patient management. Because 7-day phone follow-up was unreliable in prior site studies^16^ and infeasible to power, we expanded the simulated-patient method of Das et al^23^ to evaluate changes in patient management and assess potential harm from such changes. Field workers mimicked five common, well-defined infections for which the indication for treatment, counselling or referral is clear from treatment guidance: (i) acute gastroenteritis (young child, accompanied by mother), (ii) acute severe pneumonia (caretaker seeking care for sick older adult at home), (iii) acute fever with no other symptoms, (iv) acute urinary tract infection (UTI) in an adult woman, (v) acute pharyngitis (scenarios and scoring grids in appendix pp14). Finally, a post-intervention process evaluation consisted of focus-groups and in-depth interviews.

### Sampling strategy and sample size

#### Patient surveys

At every participating community-level healthcare provider, patients or parents of paediatric patients were surveyed consecutively after completing a healthcare visit, regardless of whether they received an antibiotic. As soon as an interviewer completed a survey, the next patient finishing a visit was enrolled. This continued over several days until 100 patient surveys were completed. Sample size calculations indicated that with 11 clusters per intervention arm per site, 100 patient surveys per provider type per village/neighbourhood cluster, a frequency of Watch-group antibiotic use of 24% (as observed previously in Kimpese^15^), an assumed intraclass correlation coefficient of 0.01, and a two-sided α of 0·05, there would be 80% power to detect a 40% net difference (i.e. (I_post - I_base) - (C_post - C_base)) in Watch-group antibiotic use. A simple difference (I_post - I_base) of >30% can be estimated with >80% power (appendix pp20).

#### Household surveys

Per village/neighbourhood, 36 households were randomly selected from the Health Demographic Surveillance Site database (updated in 2021) in Nanoro and were randomly spatially sampled in Kimpese. GPS points were randomly distributed within a polygon with outer limits of each village, and the nearest house to each point was selected. Household heads (if not available, another adult household member present at the time of the visit) were surveyed on household structure, and healthcare provider visits (including any medicine vendors) among all household members.

#### Simulated patient visits

Before each visit, field agents were trained to present at the healthcare provider and answer questions in a standardised way. Visits were unannounced. Providers consented minimum two months before baseline visits, to avoid providers altering their behaviour during visits. Post-intervention visits followed the third intervention round by up to two months.

#### Process evaluation

In five Nanoro villages, participants to focus-groups and in-depth-interviews were recruited through purposive sampling (appendix pp20). Two external evaluators conducted 2 focus-groups, each with 5 to 8 men and women from the community, and 10 in-depth-interviews with religious leaders, community leaders, and informal medicine vendors.

### Data collection and data analysis

#### Patient surveys

recorded in an electronic questionnaire reason for the visit (acute illness, chronic illness, no illness), clinical signs and symptoms, (if determined) clinical diagnosis, quantity of antibiotics dispensed by group of antibiotics if any, dose and duration of antibiotic treatment, mode of administration, and antimalarials used concomitantly. In Kimpese, interviewers recording patient surveys were medical doctors, who assigned an infection (or clinical presentation when no specific infection could be assigned) to each patient, based on clinical signs and symptoms and a potential diagnostic test result during the visit. In Nanoro, interviewers were generally nurses, who recorded diagnoses or clinical presentations from the patient’s health booklet, when available. Based on recorded diagnoses, clinical signs and symptoms, and diagnostic test results, BI and DV independently assigned an infection to each acute illness visit (appendix pp4). We excluded patient surveys at providers that were not surveyed both at baseline and endline, and at providers that had fewer than 20 surveys completed at either base- or endline.

#### Household surveys

recorded the frequency of healthcare provider visits in the previous month and previous three months of each household member, with the choice of provider. Using the person-months surveyed as denominator, we estimated the monthly rate of healthcare utilisation by provider type, in each study site. The overall rate is the sum of provider type-specific rates. The Kimpese survey did not distinguish public health centres from private clinics. To infer provider-specific rates, we therefore used the proportional distribution of visits during a 2020–21 Kimpese household survey, which reported 25·5 (95% CI 24·6–26·4) visits per 1000 inhabitants per month to public health centres and 31·0 (95% CI 30·0–32·0) to private clinics.^16^ We inferred the monthly number of provider visits to each of the providers in the study by multiplying the rate of provider-type-specific healthcare utilisation with the population size of the village/neighbourhood cluster.

We estimated the prevalence of (Watch-group) antibiotic use among patients visiting community-level providers for acute illness at baseline and post-intervention, correcting for two-stage sampling (using the survey package in R) and post-stratification weighting for the estimated number of provider visits.

We then estimated prevalence ratios (PR) of (Watch-group) antibiotic use in the intervention versus control group, using negative binomial regression with cluster-robust (sandwich) standard errors (MASS and sandwich packages in R), accounting for overdispersion and non-independence of observations within providers. Models included the number of patients using (Watch-group) antibiotics as outcome, an offset term for the log of the expected number of patients per provider (village population × provider-specific utilization rate), the intervention × time (baseline/post-intervention) interaction, and randomisation strata. The interaction term coefficient represents the difference-in-differences estimate of the intervention effect. For subgroup analyses by site, provider type and assigned infection (post-hoc), we assessed effect modification by adding a three-way intervention × time × subgroup interaction term, with Wald-test p-values reported.

In health centres and private clinics within intervention clusters, we described the proportion of antibiotic use that could have been substituted (either by no antibiotic or by an antibiotic from a different AWaRe group) if prescribers had adhered to the AWaRe Antibiotic Book’s recommended AWaRe group for each infection.

Combining the provider-type specific adjusted prevalence of (Watch-group) antibiotic use and rate of provider-type-specific healthcare utilisation, we estimated the rate of antibiotic use per 1000 inhabitants per month, by study site. For 95% confidence intervals, we combined standard errors assuming independent samples.

After simulated patient visits, field agents completed a scoring grid for each simulated infection to record history taking questions, physical examinations, counselling, medicine dispensing or referrals during the visit. Each question or action was assigned a predefined weight (positive or negative) reflecting its relative importance in patient management, as agreed upon by DV, BM, SD and BI (three clinicians, one pharmacist; from Burkina Faso, Democratic Republic of Congo, and Belgium).

Summing these weighted components produced a total patient management score (range −5 to 32) that quantitatively captured the provider’s adherence to recommendations of the WHO Antibiotic Book and WHO’s Integrated Management of Childhood Illnesses (2008 and its revision of 2014).

We described the provider-type-specific distribution of patient management scores during visits with the five simulated infections, comparing baseline and post-intervention by study arm. We estimated the intervention effect using linear regression with simulated infection, provider type, time, intervention arm, their interaction, and site. Again, the difference-in-differences effect and 95% confidence interval were derived from the interaction term coefficient.

#### Process evaluation

semi-structured interviews followed an interview guide with predefined themes: perceptions of antibiotics/AMR, sales practices, social dynamics related to interventions. These themes guided initial (manual) coding of transcripts, while emergent themes were incorporated to capture unanticipated insights, to compare convergent and divergent views across participants.

## Results

The healthcare provider intervention component enrolled 18 health centres (9 in Kimpese, 9 in Nanoro), 12 private clinics (only in Kimpese), 32 community pharmacies (22 in Kimpese, 2 in Nanoro), and 22 informal medicine vendors (only in Nanoro, figure 1). The health education campaign aimed to reach an estimated 30131 residents in Kimpese and 43933 in Nanoro.

**Figure 1.**
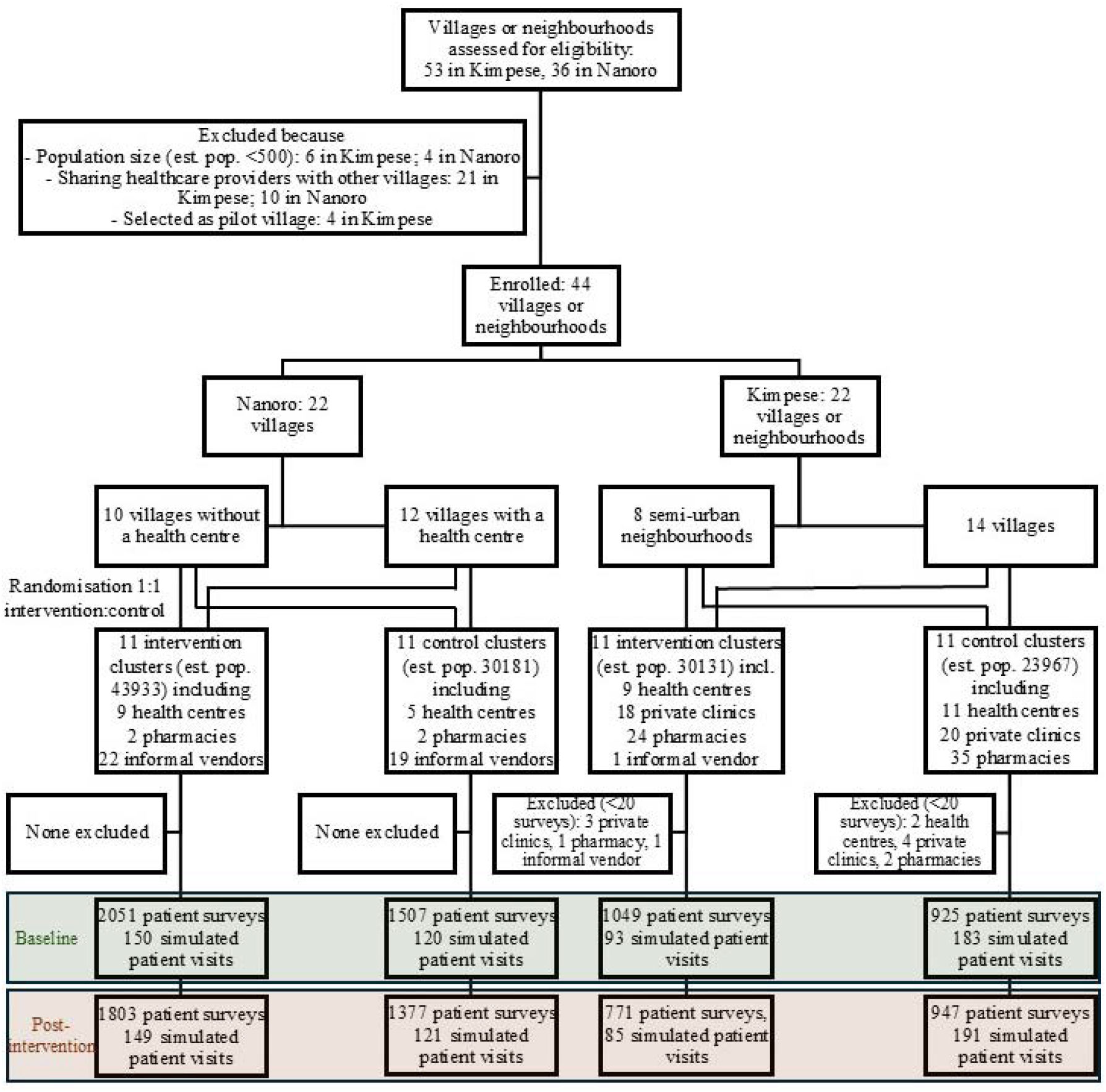
Trial profile

We excluded 13 providers with <20 patient surveys (Figure 1). At baseline, 5532 patient surveys were completed (1974 at 105 providers in Kimpese, 3558 at 59 providers in Nanoro; Oct 26, 2022, to Mar 13, 2023) and 4898 completed patient surveys post-intervention (1718 in Kimpese, 3180 in Nanoro; Nov 6, 2023 to Apr 3, 2024). Patients were surveyed at 32 health centres (18 intervention, 14 control), 31 private clinics (15 intervention, 16 control), 60 pharmacies (25 intervention, 35 control), and 41 informal vendors (22 intervention, 19 control). Malaria, acute respiratory infections, skin or soft tissue infections, and unexplained fever were the most frequent infections recorded (Table 2).

**Table 2.**
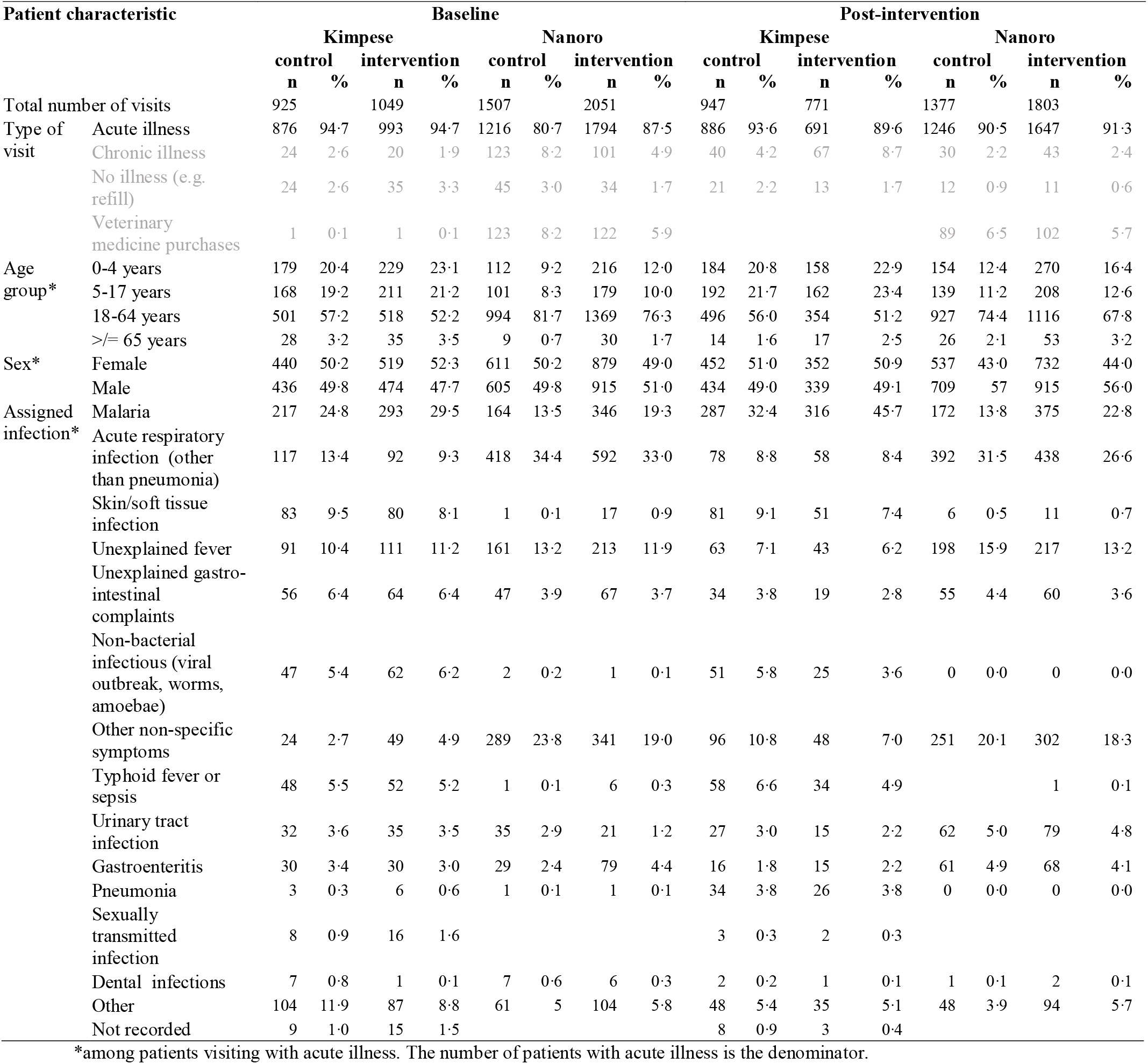
Characteristics of participants to patient surveys, Oct 2022-Feb 2023 and Oct 2023-Mar 2024.

| Patient characteristic |  | Baseline |  |  |  |  |  |  |  | Post-intervention |  |  |  |  |  |  |  |
| --- | --- | --- | --- | --- | --- | --- | --- | --- | --- | --- | --- | --- | --- | --- | --- | --- | --- |
|  |  | Kimpese |  |  |  | Nanoro |  |  |  | Kimpese |  |  |  | Nanoro |  |  |  |
|  |  | control<br>n | % | intervention<br>n | % | control<br>n | % | intervention<br>n | % | control<br>n | % | intervention<br>n | % | control<br>n | % | intervention<br>n | % |
| Total number of visits |  | 925 |  | 1049 |  | 1507 |  | 2051 |  | 947 |  | 771 |  | 1377 |  | 1803 |  |
| Type of visit | Acute illness | 876 | 94.7 | 993 | 94.7 | 1216 | 80.7 | 1794 | 87.5 | 886 | 93.6 | 691 | 89.6 | 1246 | 90.5 | 1647 | 91.3 |
|  | Chronic illness | 24 | 2.6 | 20 | 1.9 | 123 | 8.2 | 101 | 4.9 | 40 | 4.2 | 67 | 8.7 | 30 | 2.2 | 43 | 2.4 |
|  | No illness (e.g. refill) | 24 | 2.6 | 35 | 3.3 | 45 | 3.0 | 34 | 1.7 | 21 | 2.2 | 13 | 1.7 | 12 | 0.9 | 11 | 0.6 |
|  | Veterinary medicine purchases | 1 | 0.1 | 1 | 0.1 | 123 | 8.2 | 122 | 5.9 |  |  |  |  | 89 | 6.5 | 102 | 5.7 |
| Age group* | 0-4 years | 179 | 20.4 | 229 | 23.1 | 112 | 9.2 | 216 | 12.0 | 184 | 20.8 | 158 | 22.9 | 154 | 12.4 | 270 | 16.4 |
|  | 5-17 years | 168 | 19.2 | 211 | 21.2 | 101 | 8.3 | 179 | 10.0 | 192 | 21.7 | 162 | 23.4 | 139 | 11.2 | 208 | 12.6 |
|  | 18-64 years | 501 | 57.2 | 518 | 52.2 | 994 | 81.7 | 1369 | 76.3 | 496 | 56.0 | 354 | 51.2 | 927 | 74.4 | 1116 | 67.8 |
|  | >= 65 years | 28 | 3.2 | 35 | 3.5 | 9 | 0.7 | 30 | 1.7 | 14 | 1.6 | 17 | 2.5 | 26 | 2.1 | 53 | 3.2 |
| Sex* | Female | 440 | 50.2 | 519 | 52.3 | 611 | 50.2 | 879 | 49.0 | 452 | 51.0 | 352 | 50.9 | 537 | 43.0 | 732 | 44.0 |
|  | Male | 436 | 49.8 | 474 | 47.7 | 605 | 49.8 | 915 | 51.0 | 434 | 49.0 | 339 | 49.1 | 709 | 57 | 915 | 56.0 |
| Assigned infection* | Malaria | 217 | 24.8 | 293 | 29.5 | 164 | 13.5 | 346 | 19.3 | 287 | 32.4 | 316 | 45.7 | 172 | 13.8 | 375 | 22.8 |
|  | Acute respiratory infection (other than pneumonia) | 117 | 13.4 | 92 | 9.3 | 418 | 34.4 | 592 | 33.0 | 78 | 8.8 | 58 | 8.4 | 392 | 31.5 | 438 | 26.6 |
|  | Skin/soft tissue infection | 83 | 9.5 | 80 | 8.1 | 1 | 0.1 | 17 | 0.9 | 81 | 9.1 | 51 | 7.4 | 6 | 0.5 | 11 | 0.7 |
|  | Unexplained fever | 91 | 10.4 | 111 | 11.2 | 161 | 13.2 | 213 | 11.9 | 63 | 7.1 | 43 | 6.2 | 198 | 15.9 | 217 | 13.2 |
|  | Unexplained gastro-intestinal complaints | 56 | 6.4 | 64 | 6.4 | 47 | 3.9 | 67 | 3.7 | 34 | 3.8 | 19 | 2.8 | 55 | 4.4 | 60 | 3.6 |
|  | Non-bacterial infectious (viral outbreak, worms, amoebae) | 47 | 5.4 | 62 | 6.2 | 2 | 0.2 | 1 | 0.1 | 51 | 5.8 | 25 | 3.6 | 0 | 0.0 | 0 | 0.0 |
|  | Other non-specific symptoms | 24 | 2.7 | 49 | 4.9 | 289 | 23.8 | 341 | 19.0 | 96 | 10.8 | 48 | 7.0 | 251 | 20.1 | 302 | 18.3 |
|  | Typhoid fever or sepsis | 48 | 5.5 | 52 | 5.2 | 1 | 0.1 | 6 | 0.3 | 58 | 6.6 | 34 | 4.9 |  |  | 1 | 0.1 |
|  | Urinary tract infection | 32 | 3.6 | 35 | 3.5 | 35 | 2.9 | 21 | 1.2 | 27 | 3.0 | 15 | 2.2 | 62 | 5.0 | 79 | 4.8 |
|  | Gastroenteritis | 30 | 3.4 | 30 | 3.0 | 29 | 2.4 | 79 | 4.4 | 16 | 1.8 | 15 | 2.2 | 61 | 4.9 | 68 | 4.1 |
|  | Pneumonia | 3 | 0.3 | 6 | 0.6 | 1 | 0.1 | 1 | 0.1 | 34 | 3.8 | 26 | 3.8 | 0 | 0.0 | 0 | 0.0 |
|  | Sexually transmitted infection | 8 | 0.9 | 16 | 1.6 |  |  |  |  | 3 | 0.3 | 2 | 0.3 |  |  |  |  |
|  | Dental infections | 7 | 0.8 | 1 | 0.1 | 7 | 0.6 | 6 | 0.3 | 2 | 0.2 | 1 | 0.1 | 1 | 0.1 | 2 | 0.1 |
|  | Other | 104 | 11.9 | 87 | 8.8 | 61 | 5 | 104 | 5.8 | 48 | 5.4 | 35 | 5.1 | 48 | 3.9 | 94 | 5.7 |
|  | Not recorded | 9 | 1.0 | 15 | 1.5 |  |  |  |  | 8 | 0.9 | 3 | 0.4 |  |  |  |  |

\*among patients visiting with acute illness. The number of patients with acute illness is the denominator.

Healthcare visits during the past month were recorded from 5048 individuals (1326 in Kimpese, 3722 in Nanoro) from 678 households (144 in Kimpese, 534 in Nanoro). In Kimpese, 62·6 visits to health centres or private clinics (95%CI 49·1-76·1) were reported per 1000 inhabitants per month, and 26·4 to community pharmacies (95%CI 17·7-35·1; appendix pp21). In Nanoro, 26·3 visits to health centres (95%CI 21·1-31·5), 2·4 visits to pharmacies (95%CI 0·8-4·0), and 5·4 visits to informal medicine vendors (95%CI 3·0-7·7) were reported per 1000 inhabitants per month.

The prevalence ratio (PR) for use of Watch-group antibiotics was 0·33 (95%CI 0·14-0·78). The weighted prevalence of use of Watch-group antibiotics decreased from 26·8% (95%CI 8·8-45) of patients to 17·1% (95%CI 7·7-26) in the intervention group, while it increased from 13·4% (95%CI 4·8-22) to 21.2% (95%CI 8·9-34) in the control group (Figure 2). The relative reduction in Watch-group antibiotic use was comparable between sites (p=0.54), despite higher prevalence at baseline in Kimpese (41·6%) than in Nanoro (3·3%). Observed baseline ICC was 0.07 at medicine vendors and 0.09 in health centres in Kimpese, 0.01 at medicine vendors and 0.04 at health centres in Nanoro.

**Figure 2.**
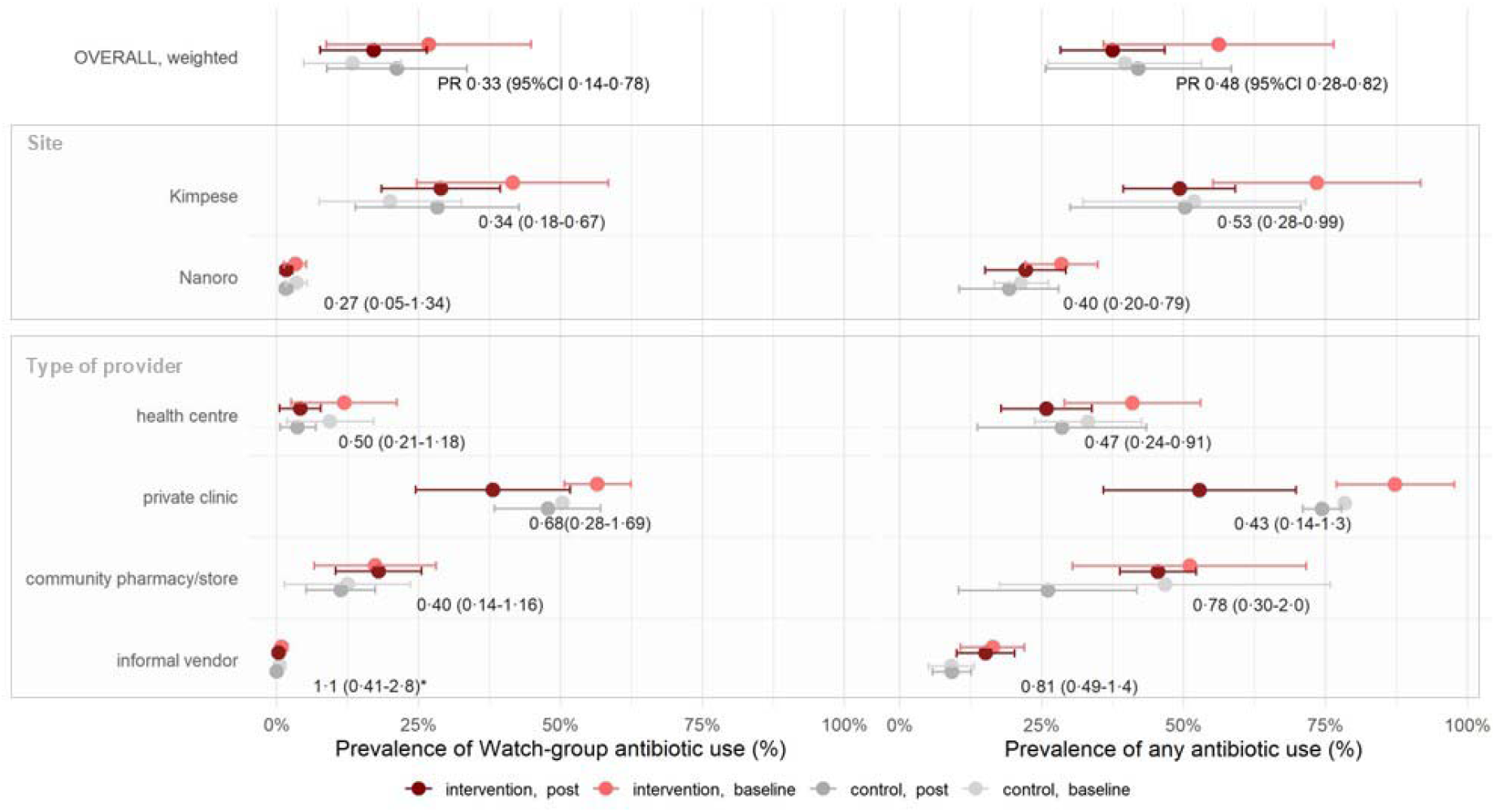
Prevalence of (Watch-group) antibiotic use during healthcare visits, overall, by site, or by type of provider, corrected for cluster surveys and offset against the expected number of visits at each provider. PR: adjusted prevalence ratio; *: in a subgroup with 0 patients with Watch-group antibiotic use, we added a small continuity correction.

PR of any antibiotic use was 0·48 (95%CI 0·28-0·82). Weighted prevalence of use of any systemic antibiotic in intervention clusters decreased from 56·2 (95%CI 35·9-76·5) to 37·5% (95%CI 28·3-46·7) while modestly increasing in control clusters (Figure 2; appendix pp23).

Exploratory analyses indicated stronger decreases in Watch-group and any antibiotic use in health centres or private clinics than at informal vendors (p_interaction_<0.01). Reductions in Watch-group antibiotic use were largest for urinary tract infections, unexplained fever, acute respiratory infections, and sexually transmitted infections (appendix pp24). Any antibiotic use decreased most in sexually transmitted infections, malaria, unexplained fever, and gastro-enteritis.

When adhering to AWaRe Antibiotic Book treatment guidance in health centres and private clinics, 413 (65·9%) antibiotic treatment episodes could have been avoided at baseline, and 185 (54·1%) post-intervention (Figure 3). Watch-group antibiotics made up 240 (38·3%) antibiotic treatment episodes at baseline, of which 199 (82·9%) could have been substituted by access antibiotics or by no antibiotic treatment. Post-intervention, 116 (33·7%) treatment courses had Watch-group antibiotics, of which 93 (80·2%) could be substituted.

**Figure 3.**
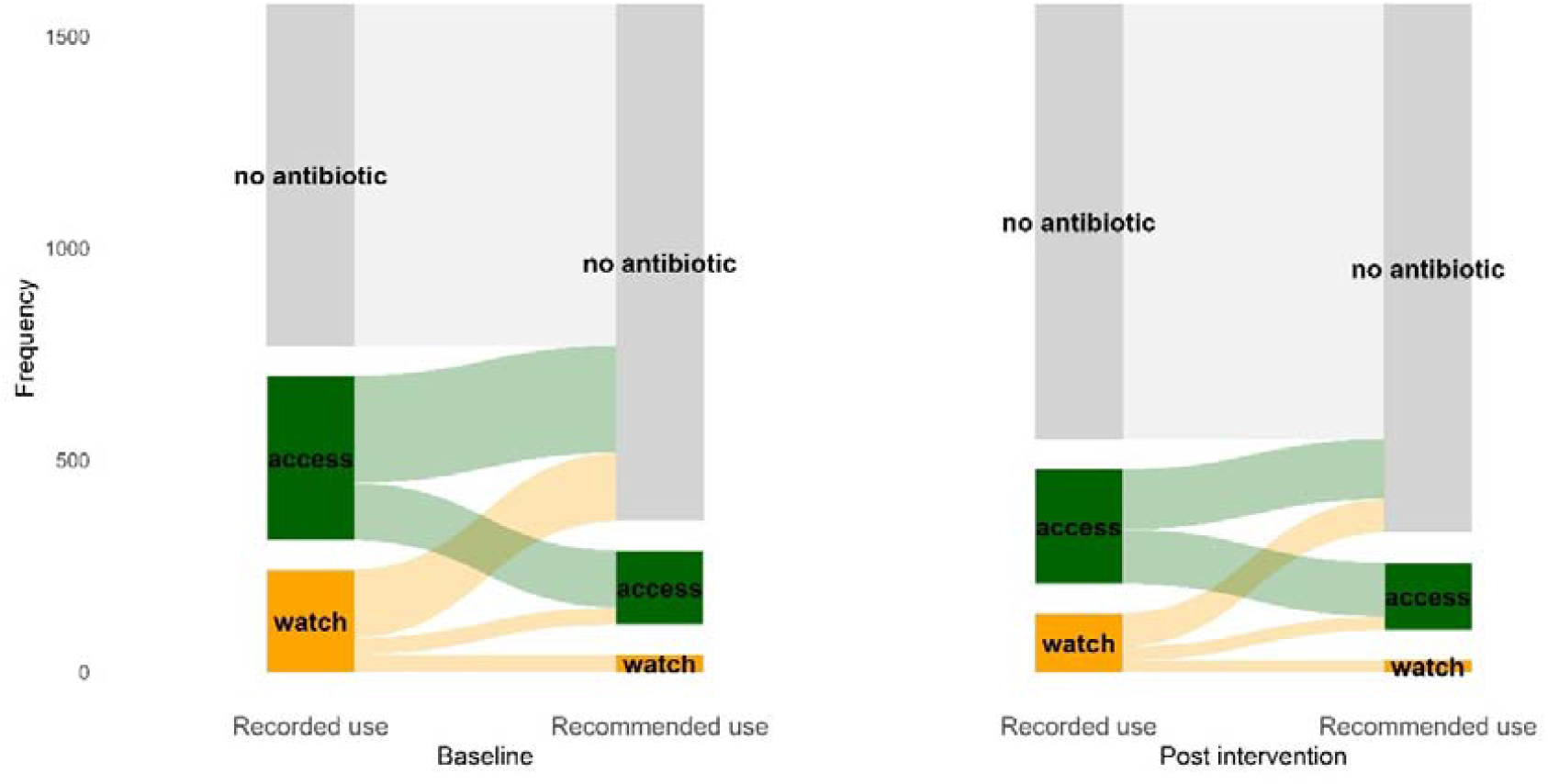
Distribution of recorded and of recommended use of Access/Watch/Reserve-group antibiotics at baseline and post-intervention in health centres and private clinics in intervention clusters. Recommended use was according to infection-specific AWaRe Book treatment guidance.

In intervention villages in Kimpese, the rate of population-wide use of any antibiotic decreased from 61·9 (95%CI 61·2-62·6) to 40·6 treatment episodes per 1000 inhabitants per month (95%CI 39·9-41·3, appendix pp25). Use of Watch-group antibiotics decreased from 33·3 (95%CI 24·2-47·5) to 23·1 episodes per 1000 inhabitants per month (95%CI 13·1-43·5). Most of the reduction in antibiotic use was at private clinics (decrease of 11·9 antibiotic treatment episodes, 95%CI 22·5-1·3; 6·3 Watch-group episodes per 1000 inhabitants per month, 95%CI-2·5-15·2) and health centres (decrease 8·0 antibiotic treatment episodes, 95%CI 0·4-16·6; 4·0 Watch-group episodes per 1000 inhabitants per month, 95%CI -6·2-14·1).

In Nanoro, use of any antibiotic decreased from 9·9 (95%CI 8·8-11·0) to 7·6 episodes per 1000 inhabitants per month (95%CI 6·5-8·7) and use of Watch-group antibiotics decreased from 1·1 (95%CI 0·0-2·3) to 0·6 per 1000 inhabitants per month (95%CI 0·0-2·6). Most of the reduction in antibiotic use was at health centres (decrease of 2·0 antibiotic treatment episodes, 95%CI 1·5-2·5; 0·5 Watch-group episodes per 1000 inhabitants per month, 95%CI-0·80 to 1·80).

We repeatedly conducted 546 simulated patient visits (Figure 1). At baseline, across providers and infections, patient management scores ranged between -4 and 24; mean 4·3 (95%CI 3·8-4·8). Scores were higher at health centres (mean 6·6, 95%CI 5·7-7·4) and community pharmacies (mean 6·3, 95%CI 5·5-7·0) than at informal medicine vendors (mean 0·1, 95%CI -0·1-0·3; Figure 4). Scores were higher at health centres (mean 9·5, 95%CI 8·0-10·9) and pharmacies in Kimpese (mean 6·7, 95%CI 5·8-7·5) than in Nanoro (respectively 4·2, 95%CI 3·5-4·9, and 2·0, 95%CI 0·88-3·2; appendix pp27).

**Figure 4.**
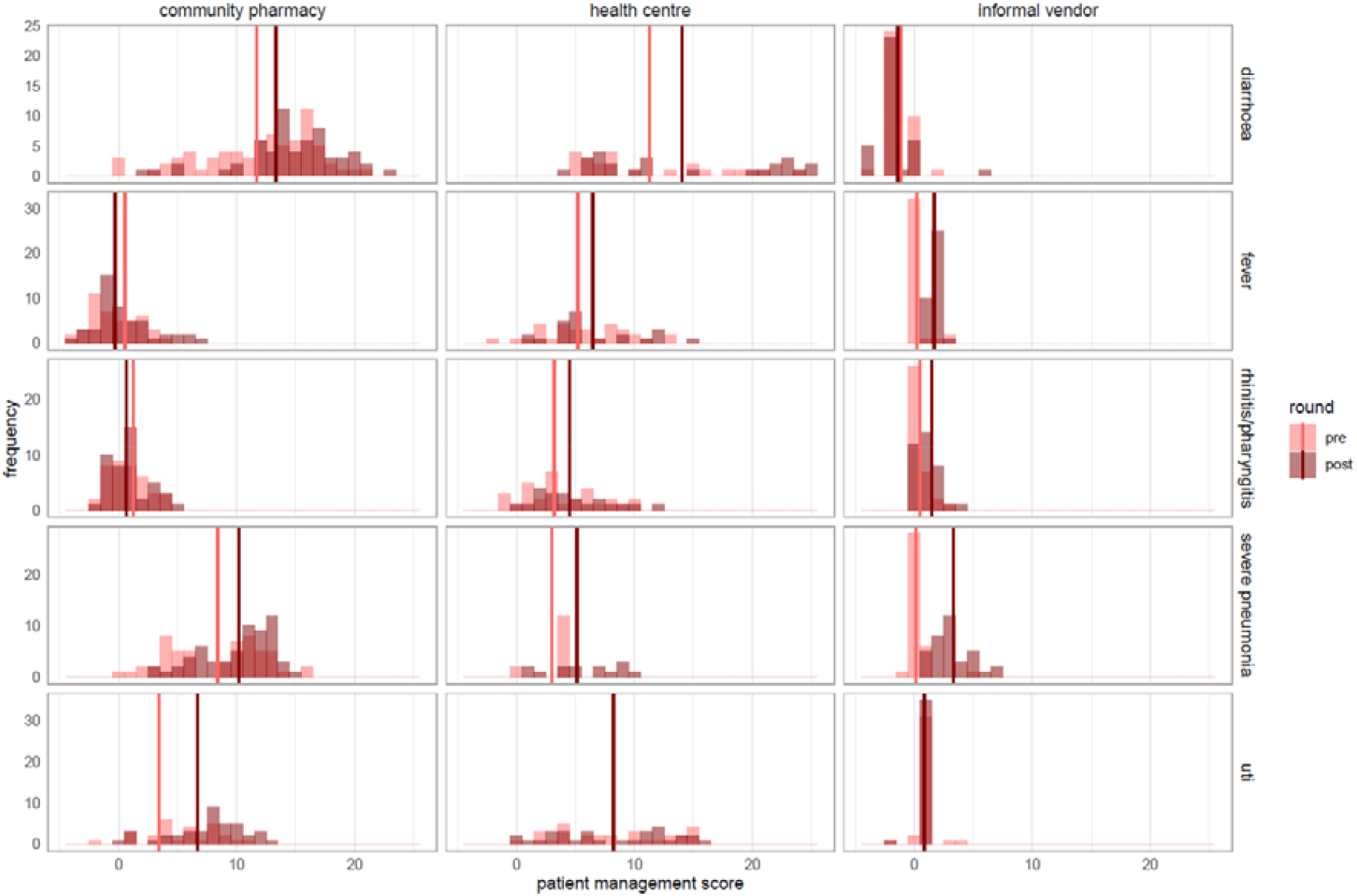
Distribution of patient management scores at all healthcare providers in the intervention group before vs after intervention, faceted by type of provider and by infection. Vertical lines are mean scores. Higher scores indicate better patient management.

Changes in patient management score were minimal. The difference-in-difference change in patient management score was a 1·5 point increase (95%CI-0·77-3·68) in health centres, 0·29 point increase (-0·21-0·78) during informal vendor visits, and a -1·0 point decrease (95%CI -2·3-0·23) at pharmacies. We did not observe major changes in use of Watch-group antibiotics during visits, except a possible increase during diarrhoea visits to pharmacies (0 to 13·3%, 95%CI 0-30·5%) and informal vendors (0 to 31·6%, 95% 10·7-52·5%, appendix pp28).

Community members and medicine vendors consistently reported that the intervention incited partial awareness on the risks of self-medication with antibiotics, possibly resulting in a shift away from informal vendors. Some interviewees were aware that *street medicines* [i.e. medicines dispensed by informal medicine vendors] *are bad for health* but did not understand how.

> *“I didn’t know that street medicines at best mask the illness and end up causing problems”-*female community member
>
> *“Informal street vendors are no longer common in [village] because people no longer buy their products”-*village leader

Interviewees argued that greater awareness of better treatment outcomes could discourage patients from reverting to self-medication. Nevertheless, financial barriers to visiting health centers remained, even with free-of-cost healthcare for <5 year-olds. Informal vendors offered more financial flexibility.

> *“Abandoning makeshift pharmacies in favour of health centres is possible because differences [in clinical outcome] motivated people[*…*]if treatment is more effective at the health centre, those who switched will continue to do so”-*female community member
>
> *“There are no more medicines at the hospital. The government said to treat children for free, and the health centres go bankrupt. They prescribe and tell you to go to the [community] pharmacy. You spend 1000CFA-francs on gasoline. When you arrive, they ask for 3000CFA-francs[*…*]Informal vendors pharmacies gained power because of this. Even if it means buying 100CFA-francs products and dying, we’ll do it, because we have no other choice”*-male community member

Informal vendors highlighted they faced pressure to sell antibiotics clients asked for. Refusing to sell antibiotics or patient referrals resulted in client dissatisfaction.

> *“Customers usually insist on the medications they want, while others refuse to follow[*…*]advice. As a result, customers chose to buy elsewhere”-*informal vendor
>
> *“On a daily basis, 20% of customers get angry and leave when I refer them to the health centre”-*informal vendor
>
> *“Patients expect to receive medication at every visit, and this puts [vendors] in difficult situations”*-informal vendor

## Discussion

A co-created intervention bundle that combined repeated educational and feedback sessions targeting all community-level healthcare providers with an AMR awareness campaign in surrounding communities, was feasible and effective in reducing (Watch-group) antibiotic use. Patient management changed slightly, indicating no harm and potentially modest improvement in health centres.

Across provider types, the intervention achieved a substantial relative reduction in Watch-group- antibiotic use compared to the concurrent increase in the control group (PR 0·33, 95%CI 0·14-0·78), the primary outcome. This finding is consistent with preparatory studies indicating that >75% of Watch-group use could be substituted by Access antibiotics or no antibiotic.^15,17^ Antibiotic use was reduced by half, an unexpected result given that the intervention focused on Watch-group antibiotics and that dispensing no antibiotic could go against patients’ expectations.

Exploratory analyses suggested that a shift to no antibiotic treatment might have been more pronounced in health centres and private clinics than at medicine vendors, and for infections that were prioritized in the provider intervention. In Nanoro, the change in per capita antibiotic use was attributable to reduced use in health centres. In Kimpese, reductions in private clinics contributed at least as much as those in health centres, consistent with prior studies indicating that half of community-wide antibiotic use stemmed from visits to private providers.^16^ Quantitative and qualitative findings confirm medicine vendors’ difficulties in reducing over-the-counter dispensing of (Watch-group) antibiotics. Educational sessions to informal vendors in India also showed no change in the prevalence of antibiotic use, but improved patient management.^23^ Informal vendors highlighted difficulties in influencing antibiotic use resulting from patient expectations. The intervention intended to improve awareness among both providers and general population, aiming at a synergetic effect with provider training to rationalise supply and demand. Formal and informal medicine vendors are also visited because of financial, (waiting) time, antibiotic availability, and proximity considerations.^13,24^ Certain policies, such as free-of-charge healthcare for young children, strengthened patient-centred care, or regulation of antibiotic sales have proven successful in curbing self-medication.^24,25^

Educational and feedback sessions with communities, prescribers and dispensers were intended to enable integration in routine supervision activities by health district staff. The intervention therefore focused on infections with the largest potential antibiotic use improvements, identified in prior surveys, and integrated existing treatment guidance based on Integrated Management of Childhood Illness with the AWaRe Antibiotic Book, published months before the intervention start. Even without (costly) structural or restrictive interventions, community-wide antibiotic use can be substantially optimized with limited (human) resources, helping to curb antimicrobial resistance, reduce unnecessary healthcare expenditure, and prevent adverse events.

Because unintended effects were possible, we used simulated patient visits to monitor care quality and safety. Changes in clinical outcome following primary-care or pharmacy visits are difficult to detect – often underpowered with challenging follow-up. Simulated patient visits detected no worsening of patient management, instead a modest improvement, mostly in primary care clinics. The extent to which clinical history was taken, examinations were performed, referrals made, and treatments provided varied widely across sites and provider types. Informal vendors rarely took an anamnesis and primarily dispensed medicines, with little or no change after the intervention.

Despite marked reductions, most Watch-group antibiotic use remained outside AWaRe Book recommendations. With low baseline performance and modest improvements in patient management, substantial potential remains for strengthening community-based antimicrobial stewardship.

Adjusting antibiotic-use prevalence for healthcare utilisation and village population size allowed estimation of population-wide antibiotic use, highlighting the intervention’s public health implications; however, because utilisation offsets and survey weights were partly estimated from self-reporting rather than directly observed, measurement error could have added uncertainty to the final estimates. Our study did not measure changes in healthcare utilisation. Community feedback suggested a shift from self-medication to health-centre consultations, which – had it been incorporated into the weighting – might have yielded a more accurate estimate of post-intervention antibiotic use and may mean our current estimate potentially underestimates the effect on population-wide antibiotic use. Another limitation was that we could not formally power our study for the presented sub-analyses by infection or provider type due to a limit in eligible villages and providers. Furthermore, lower than anticipated Watch-group antibiotic use in Nanoro, differences in healthcare seeking, and higher clustering impacted the precision of sub-group analyses and limited interpretability of risk differences. However, the consistency in the direction of effect and relative reduction in antibiotic use across settings supports the validity of the results. Some infections may have been wrongly assigned, particularly in Nanoro where surveyors had no medical background, potentially attenuating infection-specific effects. Finally, per capita antibiotic use rates translate prevalence estimates to population-wide impact of the intervention, but did not consider doses or duration of treatment. Correct antibiotic use is also impacted by dose, actual duration, adherence and interruption of treatment, which we could not reliably assess from patient surveys.

## Conclusion

A contextualised behavioural intervention targeting both supply (medicine vendors and primary care) and demand (communities), focusing on key, common primary-care infections, was found to be feasible and resulted in decreased antibiotic use, overall and of Watch-group antibiotics. Reductions in antibiotic use were greatest in clinics and health centres, with private clinics contributing importantly to the reduction. Reducing antibiotic sales from medicine vendors proved difficult with only educational and feedback interventions. The bundle did not negatively impact patient management, indicating no harm. Reduced community-level use of broad-spectrum antibiotics could help slow community-acquired pathogens’ increasing resistance to clinically important antibiotics.

## Data Availability

Questionnaires, informed consent forms, pseudonymized datasets, data dictionaries, and data analysis scripts in R are available on a Github repository

https://github.com/ingelbeen/cabu_intervention

## Contributors

BI, DV, EvK, EW, BC, DMP and MvdS conceptualised the study; BI, DV and BM developed and validated questionnaires with input from LC and SD; BI, DV and SD developed the simulated patient visit scenarios and checklists; LC, SJK, CMKM, and MM led intervention development; BI, DV and BM coordinated data collection; DV and SJK coordinated the process evaluation; BI and DV curated, validated and analysed the data, with input from VB and EvK; BI wrote the draft manuscript with input from DV; all authors revised the first and subsequent drafts of the manuscript; all authors had full access to the study data and had final responsibility for the decision to submit for publication.

## Data sharing

Questionnaires, informed consent forms, intervention manual and material, de-identified, pseudonymized datasets, data dictionaries, and data analysis scripts in R are available: https://github.com/ingelbeen/cabu_intervention.

The study protocol was published: https://doi.org/10.1186/s13063-023-07856-2

## Acknowledgements

The study was funded under the Joint Programme Initiative AMR (grant JPIAMR2021-053) with member state co-funding by the Research Foundation Flanders (FWO), Sida, and the Medical Research Council. A framework agreement between Institute of Tropical Medicine, Clinical Research Unit of Nanoro and Centre de Recherche en Santé de Kimpese is financially supported by the Belgium Development Cooperation. BI is supported by a postdoctoral fellowship from the FWO (grant number 12A1Z25N). We thank all staff members of the Clinical Research Unit of Nanoro and the Centre de Recherche en Santé de Kimpese, healthcare providers and community leaders in Nanoro and Kimpese for their contributions to the intervention and study. We specifically thank Ardjima Naba and W. Athanase W. Oumsaoré for facilitating process evaluation interviews, and Tom Smekens for a review of statistical methods.

