## appendix for "Effect of a community-based behavioural intervention bundle to improve antibiotic use and patient management in Burkina Faso and DR Congo: a cluster randomised controlled trial"

https://doi.org/10.64898/2025.12.15.25342146

### Table of content

### Study setting and population

The study was conducted in two health districts with populations followed up within Health demographic surveillance sites (HDSS) in Nanoro district, Burkina Faso, and Kimpese district, DR Congo. District referral hospitals in each of both sites have a clinical microbiology laboratory and serve as an AMR surveillance site. Both research institutes coordinating the HDSS also had experienced social science teams prior to the study.

The HDSS in Nanoro was established in 2009. We screened the Nanoro HDSS area (about the third of the district surface) and its HDSS extension (another third of the surface) for potential study villages (at least 500 inhabitants, and at least one community-level or primary care provider functioning as the main medicine dispenser of the village population). The entire area, including Nanoro village, is rural. We selected 22 study villages in the health district, which has a total estimated population of 80203 (2023). All villages had at least one medicine vendor (pharmacy or medicine store), and 13 had a primary health centre. To ensure an even distribution of the number of patient surveys and comparable patient populations between intervention and control group, cluster villages were divided in two strata prior to randomisation. To make a random selection of households for enrolment in the household survey, a validated and up-to-date sampling frame, i.e. a list of all households participating in the HDSS with their composition and geographical coordinates, was available.

The HDSS in Kimpese was established in 2018. Kimpese health zone consists of a peri-urban town, Kimpese cité, and rural villages, with a total estimated population of 54684 (2023). Following screening of villages and town neighbourhoods, we selected 14 villages and 8 urban neighbourhoods, using the same selection criteria as in Nanoro. As in Nanoro, clusters were grouped in two strata before randomising, to ensure an even distribution of the number of patient surveys and of patient populations, although in Kimpese, strata were based on rural/village clusters and peri-urban/neighbourhood clusters. No validated up-to-date household sampling frame was available; we therefore used spatial sampling to select households in each cluster.

In Kimpese, healthcare was sought from multiple community-level and primary provider types, and rarely in the hospital: primary health centres, private clinics, and community pharmacies. What we labelled community pharmacies comprised both licensed community pharmacies with qualified dispensers, as well as medicine stores without qualified staff, functioning in a regulatory grey zone. Prior qualitative work with community members and health providers also identified traditional healers and religious leaders as occasionally consulted when seeking healthcare. Household surveys quantified the frequency each of these providers were visited (rates provided in Table S1), highlighting that two thirds of visits were to private clinics and primary health centres. Private clinics accounting for the largest fraction of visits in peri-urban areas and primary health centres more prominent in rural settings. Antibiotics were frequently dispensed across all provider types, including medicine stores, with private providers contributing disproportionately (>50% of defined daily doses per 1000 inhabitants per day) to community antibiotic use. While formal health facilities and clinics provided antibiotics within clinical consultations, medicine stores and private providers served as common points of antibiotic access outside strict prescription channels, implicating both qualified and informal sources in community antibiotic dispensing^[1]^.

In Nanoro, prior mixed methods work similarly documented a diverse healthcare-seeking environment. Communities differentiated between ‘natural’ illnesses and those thought to be magico-religious in origin, influencing where care was sought. For perceived natural illnesses, people frequently sought care at formal healthcare facilities, community pharmacies, and informal medicine vendors. For illnesses considered magico-religious, traditional healers were the primary source of care. Household surveys revealed that a about 20% of care was sought outside formal health facilities (rates provided in Table S1). This pattern was influenced by socioeconomic and access barriers such as financial limitations, proximity to informal drug vendors, long waiting times at facilities, and perceptions of healthcare workers’ attitudes^[2]^. Antibiotics in the community were often seen as general symptom relief, facilitating self-medication and use through non-clinical channels. In addition, qualitative findings from related work in the same rural setting indicate that over-the-counter antibiotic dispensing without prescription is common in both licensed pharmacies and non-licensed retail outlets, increasing opportunities for inappropriate antibiotic access and use outside formal care pathways.

Both contexts illustrate that pre-existing healthcare-seeking practices involved significant community-level antibiotic access outside formal healthcare delivery, which was considered when developing the intervention bundle.

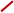

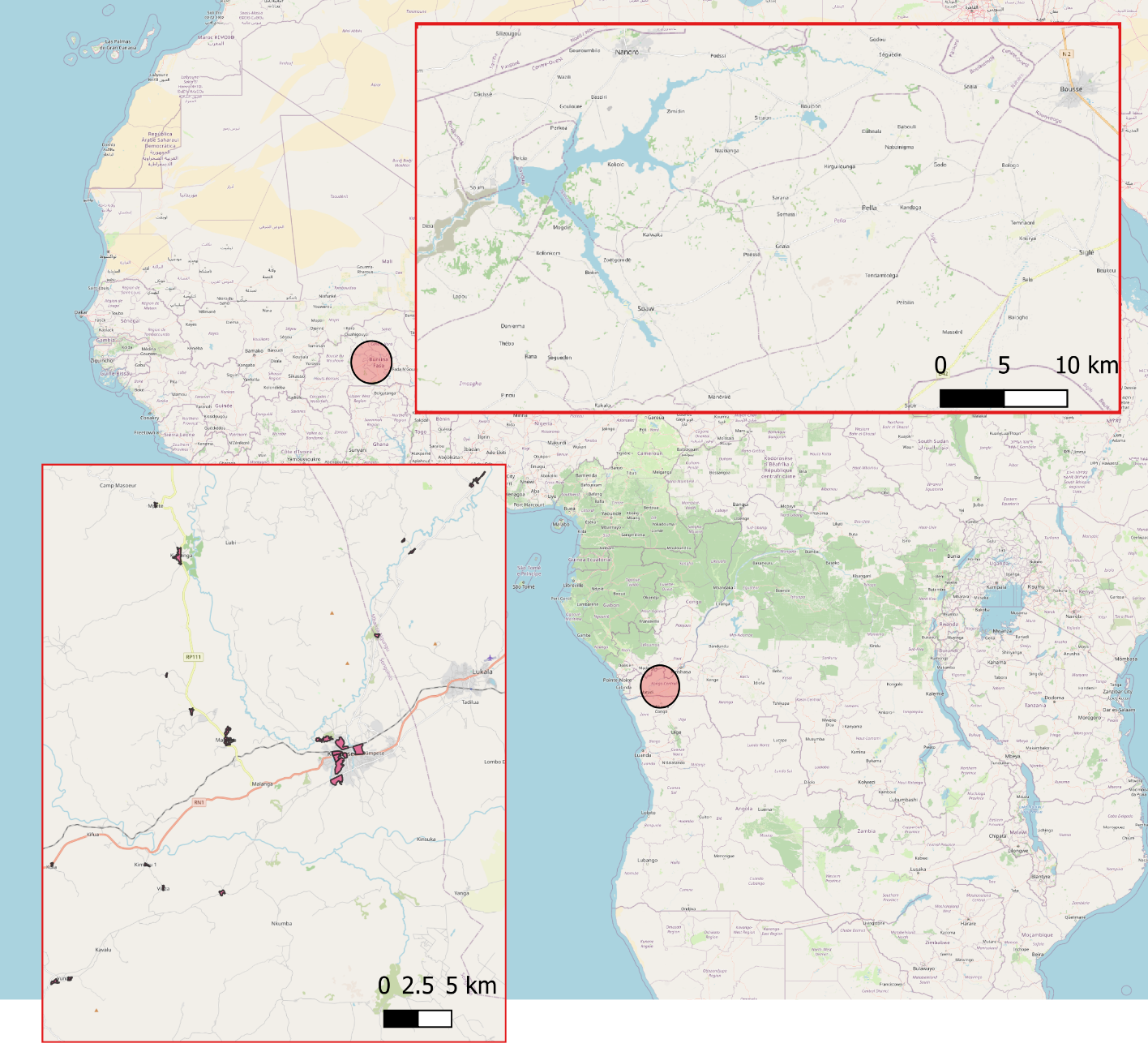

### Figure S1. Study sites in DR Congo and Burkina Faso

*Assigning infections based on reported diagnoses, clinical signs and symptoms*

Based on recorded diagnoses, clinical signs and symptoms, and diagnostic test results, two researchers independently assigned an infection to each acute illness visit, according to criteria in the Diagnosis sections in the WHO AWaRe Antibiotic Book. In case multiple infections could be assigned to the recorded clinical signs and symptoms (e.g., fever with bronchitis and a positive malaria diagnostic test), we assigned the infection that had the highest likelihood to result in antibiotic use (e.g., bronchitis). In cases of disagreement, consensus was reached through joint review. Because for certain common infections in the AWaRe Antibiotic Book, recorded clinical signs and symptoms are not specific enough to assign the specific infection, we categorised some infections in broader syndromic clinical presentations with the same treatment indications in the WHO AWaRe Antibiotic Book (e.g., bronchitis, sinusitis, pharyngitis, rhinitis and bronchiolitis grouped together as ‘acute respiratory infection (other than pneumonia)’, table below).

| **Infections in Primary Care section of WHO AWaRe Book** | **Broader infection assigned** |
| --- | --- |
| Bronchitis | Acute respiratory infection (other than pneumonia) |
| Acute otitis media | Acute respiratory infection (other than pneumonia) |
| Pharyngitis | Acute respiratory infection (other than pneumonia) |
| Acute sinusitis | Acute respiratory infection (other than pneumonia) |
| Oral and dental infections | Dental infections |
| Localized acute bacterial lymphadenitis | Other |
| Bacterial eye infections (excluding trachoma) | Other |
| Trachoma | Other |
| Community-acquired pneumonia (mild) | Pneumonia |
| Exacerbation of chronic obstructive pulmonary disease (COPD) | Acute respiratory infection (other than pneumonia) |
| Acute infectious diarrhoea & gastroenteritis | Gastroenteritis |
| Enteric fever | Typhoid fever or sepsis |
| Skin and soft tissue infections (impetigo, erysipelas, cellulitis) | Skin/soft tissue infection |
| Burn wound–related infections | Skin/soft tissue infection |
| Wound and bite–related infections | Skin/soft tissue infection |
| Chlamydial urogenital infection | Sexually transmitted infection |
| Gonococcal infection | Sexually transmitted infection |
| Syphilis | Sexually transmitted infection |
| Trichomoniasis | Sexually transmitted infection |
| Lower urinary tract infection | Urinary tract infection |
|  | Unexplained fever |
|  | Unexplained gastro-intestinal complaints |
|  | Malaria |
|  | Non-bacterial infectious (viral outbreak, worms, amoebae) |
|  | Other non-specific symptoms |

For the Nanoro patient survey data, independent assignment of infections to 6744 surveyed patients by BI and DV, resulted in 433 (6.42%) nonmatching infections. To those, all data was reviewed to reach consensus on the assigned infection.

### Supplementary text 1. Intervention manual and provider treatment guidance for prioritized infections, adapted from the 2022 AWaRe Antibiotic Book,.

Full intervention manual (original in French): <https://github.com/ingelbeen/cabu_intervention/blob/9b0cf1183b37abeddc9718fdd7ceb753e287f6d9/Guide%20d'intervention%202023%20fr.docx>

Translated intervention manual (English): <https://github.com/ingelbeen/cabu_intervention/blob/9b0cf1183b37abeddc9718fdd7ceb753e287f6d9/Intervention%20manual%202023%20en.docx>

Treatment guidance adapted from the 2022 AWaRe Antibiotic Book and IMCI (otiginal in Franch): <https://github.com/ingelbeen/cabu_intervention/blob/9b0cf1183b37abeddc9718fdd7ceb753e287f6d9/provider_intervention_guide_fr>

Modifications made as compared to the 2022 AWaRe Antibiotic Book:

- Translated from English to French (no official translation available at the time)
- Combined guidance for adults and children, because prescribers and dispensers generally dealt with all ages, and they have already a large number of guidance documents, so there was a need to integrate to the extent possible
- Removed diagnostic tests which were not available (e.g. influenza testing) or indicated as “not needed” in the AWaRe Book
- Summarised the list of “most likely pathogens”
- Selected 4 infections, making some modifications in the infections since they would not always be recognised or diagnosed as such by nurses in health centres or medicine dispensers:
  - Bronchitis was extended to cough persisting for more than five days. Bronchitis is not always diagnosed as such by most nurses
  - For community-acquired pneumonia, we aligned the CURB-65 severity scoring system with existing recognition of signs of severity: presence of 2 or more listed signs and symptoms indicated severe pneumonia and therefore referral is needed.
  - Acute diarrhoea or gastroenteritis. We removed the section on “cholera Antibiotic Treatment”
  - Acute fever without other signs: integrating existing malaria and Integrated Management of Childrhood Illness guidance. It therefore has a decision tree to check for malaria, danger signs, acute otitis media, pharyngitis, and lower urinary tract infection. Treatment for the latter three was in line with the AWaRe Book guidance.
- Removed treatment choices that were not recommended in the national essential medicines list, or not in line with existing guidance:
  - Phenoxymethylpenicillin (as potassium) 500 mg (800 000 IU) q6h ORAL as a first choice option for mild/moderate community-acquired pneumonia
  - Amoxicillin+clavulanic acid 1 g+200 mg q8h or q6h IV as a second choice in adult severe cases of community-acquired pneumonia with CURB-65 score ≥2
  - Ampicillin 50 mg/kg/dose IV/IM • ≤1wk of life: q12h • >1wk of life: q8h or Benzylpenicillin 30 mg/kg/dose (50 000 IU/kg/dose) q8h IV as potential first choice options in severe cases of community-acquired pneumonia in children
  - Sulfamethoxazole+trimethoprim 40 mg/kg SMX+8 mg/kgTMP q8h in severe cases of community-acquired pneumonia in children < 1 year and HIC positive, to treat potential Pneumocystis jirovecii pneumonia
  - Second Choice options in severe cases of community-acquired pneumonia in children if no clinical response after 48-72h: should have been referred to the hospital

Translated treatment guidance (English, note that the guidance was implemented in French, translated and adapted from the English AWaRe Book; discrepancies between the original English AWaRe Book and this guidance are possible because of the translation to French and back to english):

Treatment guidance educational material for healthcare providers

| 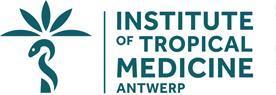 | 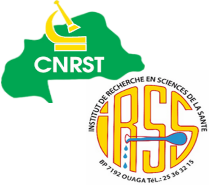 | 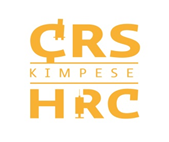 |
| --- | --- | --- |

| Study | CABU-B/C study (Optimising antibiotic use and infection control at community level through a package of behaviour-targeting interventions in Burkina Faso and DR Congo, NCT05378880) |
| --- | --- |
| Sponsor | Institute of Tropical Medicine, Nationalestraat 155, 2000 Antwerp – Belgium |
| Authors | Brecht Ingelbeen (ITM),  Valia Daniel (CRUN), |
| Material adapted from | World Health Organisation (WHO). The WHO AWaRe Antibiotic Book. Geneva; 2022 [cited 2022 Dec 12]. Available from: https://apps.who.int/iris/handle/10665/365237 |
| Version | v1.0, 7 February 2023 |

*Rationale*

The CABU-EICO project's package of interventions aims to improve the use of antibiotics, particularly Watch-group antibiotics, among dispensers (those offering antibiotics) and the general population (those requesting antibiotics). Antibiotics have been grouped into three AWaRe groups: Access = antibiotics to treat the most common infections; Watch = antibiotics that are more threatened by increasing antibiotic resistance and increase the personal risk of persistent infection after treatment, but are crucial in the treatment of certain specific bacterial infections, such as typhoid fever or other salmonellosis, sepsis, and certain sexually transmitted diseases; Reserve = last-resort antibiotics available in tertiary hospitals

The WHO AWaRe Antibiotic Guide provides recommendations on how to treat common infections in primary care, with a choice of antibiotics (or advice on when not to use antibiotics), route of administration, duration, and dose according to age.

**Table 1. Points to consider during the medical consultation**

| Diagnose | What is the clinical diagnosis?  Are there signs of a serious bacterial infection? |
| --- | --- |
| Decide | Are antibiotics really necessary?  Should I perform rapid diagnostic tests? |
| Choice of antibiotic | Which antibiotic should I prescribe?  Is it an Access, Watch or Reserve antibiotic?  Are there any allergies, interactions or other contraindications? |
| Dosage | What dose, how many times a day?  Should the dose be adjusted, for example in cases of renal impairment? |
| Administration | Which formulation should be used?  Is this a good quality product? |
| Duration | For how long?  What is the date and day of the week when treatment will be stopped? |
| Discuss | Inform the patient of the   - diagnosis - the likely duration of symptoms - the likely toxicity of the medication - what to do if they do not recover |
| Document | Record all diagnoses, decisions and management plans |

**Most patients with common benign infections can be treated without antibiotics**, as these infections are most often parasitic (such as malaria) or viral (meaning antibiotics have no effect). Antibiotic treatment increases the risk of contracting a resistant bacterial infection after treatment or other side effects. At the same time, it is essential to note that **we must not miss an opportunity to prescribe antibiotics to patients who need them** in low-resource settings, where access to care and antimicrobial treatment is often limited.

Patients who have received purely symptomatic treatment (no antimicrobials) should be clearly informed of the warning signs they should watch for and what to do if they occur.

Reducing the inappropriate use of Watch-group antibiotics is key to controlling antibiotic resistance. When antibiotics are used in primary care, the goal is for more than 90% to be Access antibiotics.

**Table 2. Infections frequently seen in primary care and recommended treatment choices**

| **Infection** | **ACCESS/ WATCH** | **First-choice antibiotic option (when an antibiotic is indicated)*** |
| --- | --- | --- |
| Bronchitis | No antibiotics | No antibiotics |
| Community-acquired pneumonia (mild cases) | ACCESS | Amoxicillin  (OR Phenoxymethylpenicillin) |
| Exacerbation of chronic obstructive pulmonary disease | No antibiotics or ACCESS | No antibiotics (for most mild cases, symptomatic treatment is the first choice and antibiotics are not necessary) OR  Amoxicillin |
| Dental infections | No antibiotics or ACCESS | No antibiotics (for most cases, the first choice is dental intervention and antibiotics are not necessary) OR  Amoxicillin OR  Phenoxymethylpenicillin |
| Infectious diarrhoea | No antibiotics or WATCH | Most mild, non-bloody diarrhoea is caused by viral infections for which antibiotics are not necessary.  In cases of **severe, acute bloody diarrhoea/dysentery** - Ciprofloxacin |
| Otitis media | No antibiotics or ACCESS | No antibiotics (most mild cases treated symptomatically) OR  Amoxicillin |
| Pharyngitis | No antibiotics or ACCESS | No antibiotics (most mild cases treated symptomatically) OR  Amoxicillin  (OR Phenoxymethylpenicillin) |
| Sinusitis | No antibiotics or ACCESS | No antibiotics (most mild cases treated symptomatically) OR  Amoxicillin OR Amoxicillin + clavulanic acid |
| Skin and soft tissue infection (mild cases) | ACCESS | Amoxicillin + clavulanic acid  OR Cloxacillin  (OR Cefalexin) |
| Lower urinary tract infection | ACCESS | Amoxicillin + clavulanic acid  (OR Sulfamethoxazole+trimethoprim  OR Nitrofurantoin  OR Trimethoprim) |

*Only oral antibiotic options are reported and recommended here.

An analysis of antibiotic use by clinical presentation showed that, if the recommendations of the AWaRe Antibiotic Guide are properly applied, in Nanoro (Burkina Faso) 69% of Watch antibiotic use could be replaced by Access antibiotic use or by no antibiotic use. In Kisantu (DR Congo), 75% of Watch antibiotics could be replaced. This would meet the WHO target of **90% of antibiotics used in primary care being from the Access group**.

The clinical presentations most frequently leading to Watch antibiotic treatments:

1. Malaria (40% of total Watch antibiotic use), even when a diagnostic test has been performed
2. Diarrhoea (14%) and other gastrointestinal complaints (10%)
3. Bronchitis (9%)
4. Skin infections (9%)

Among these clinical presentations, **only bloody diarrhoea or dysentery would require ciprofloxacin**, a Watch antibiotic. Unless the bloody diarrhoea is due to amoebiasis, which may also contain traces of blood but is less liquid (more pasty) than bacterial diarrhoea.

During **an intervention targeting prescribers and dispensers** of Nanoro in Burkina Faso and Kimpese, DR Congo, **the WHO AWaRe Antibiotic Guide**, available since December 2022, will be introduced. Below, we will limit ourselves to the clinical presentations on which the first three months of the intervention will focus. The overall objective of the intervention is to **improve the quality of care** and **optimise the use of antibiotics**, in particular to minimise the use of Watch antibiotics.

*The objective of this guide is to introduce the treatment guide for bronchitis vs. pneumonia, diarrhoea or other gastrointestinal complaints, and undifferentiated fever.*

*References*

1. World Health Organisation (WHO). The WHO AWaRe (access, watch, reserve) antibiotic book [Internet]. Geneva; 2022 [cited 2022 Dec 12]. Available from: <https://apps.who.int/iris/handle/10665/365237>

2. Keitel K, Kagoro F, Samaka J, Masimba J, Said Z, Temba H, et al. A novel electronic algorithm using host biomarker point-of-care tests for the management of febrile illnesses in Tanzanian children (e-POCT): A randomised, controlled non-inferiority trial. Tumwine JK, editor. PLoS Med [Internet]. 23 October 2017;14(10):e1002411. Available from: [https://dx.plos.org/10.1371/journal.pmed.10024](https://dx.plos.org/10.1371/journal.pmed.1002411)

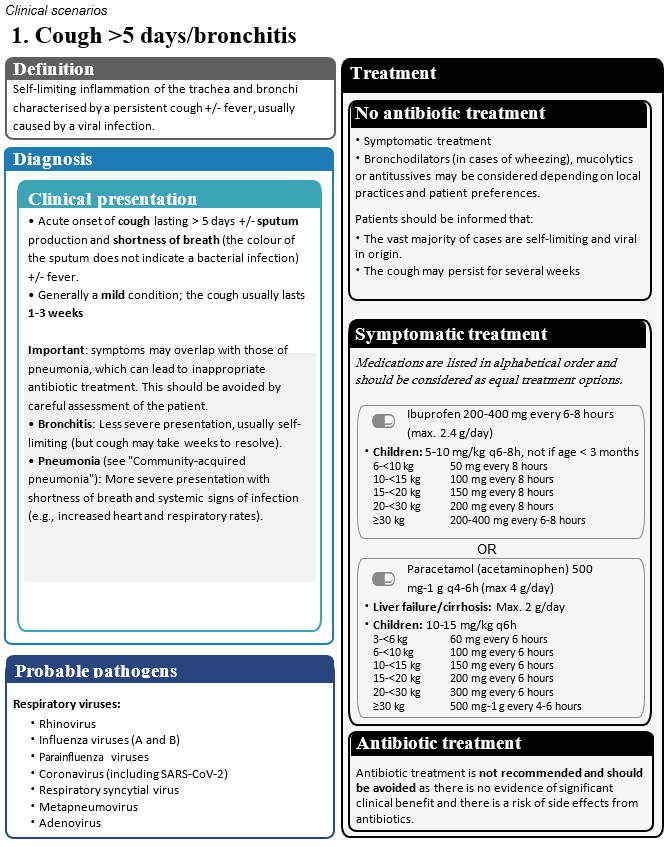

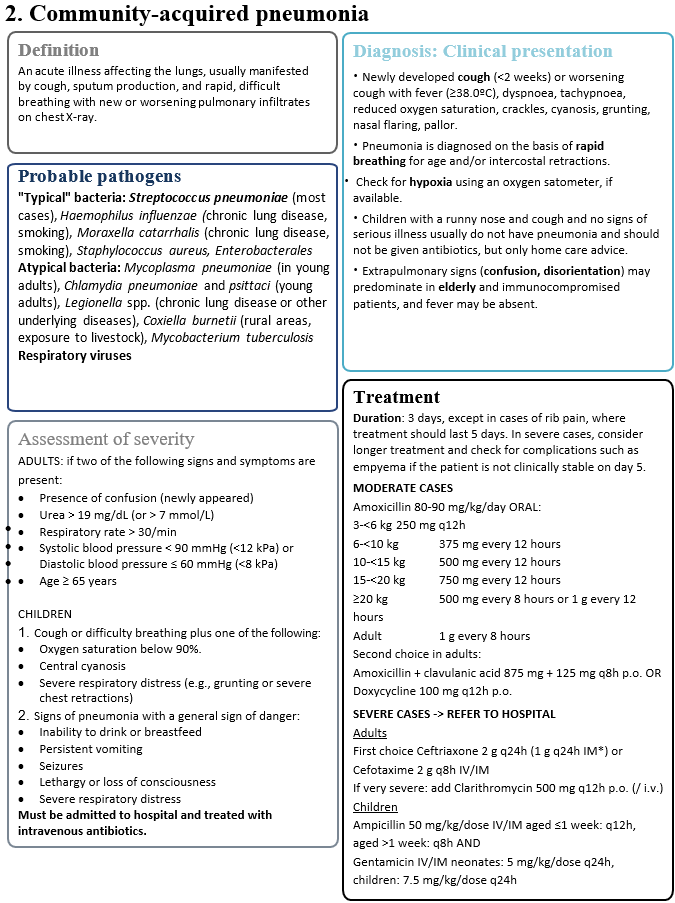

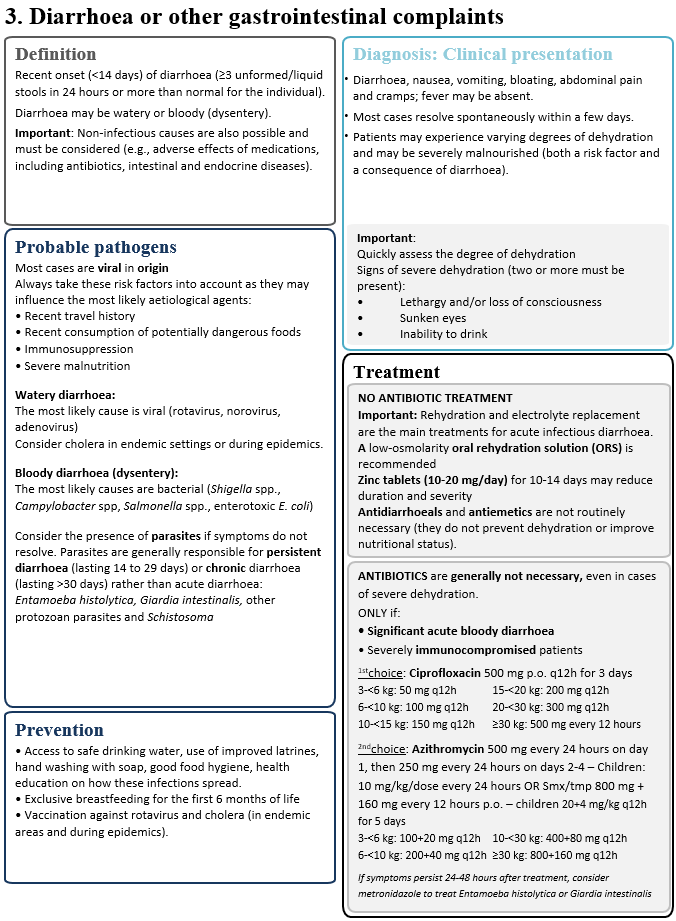

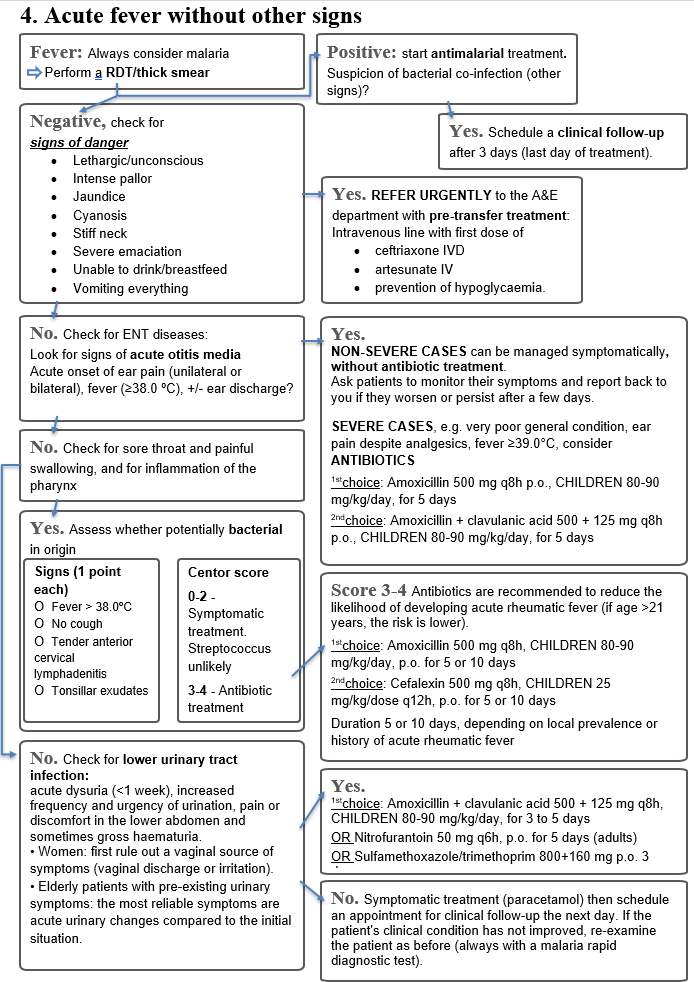

If a bacterial infection is nevertheless suspected, start an ACCESS antibiotic.

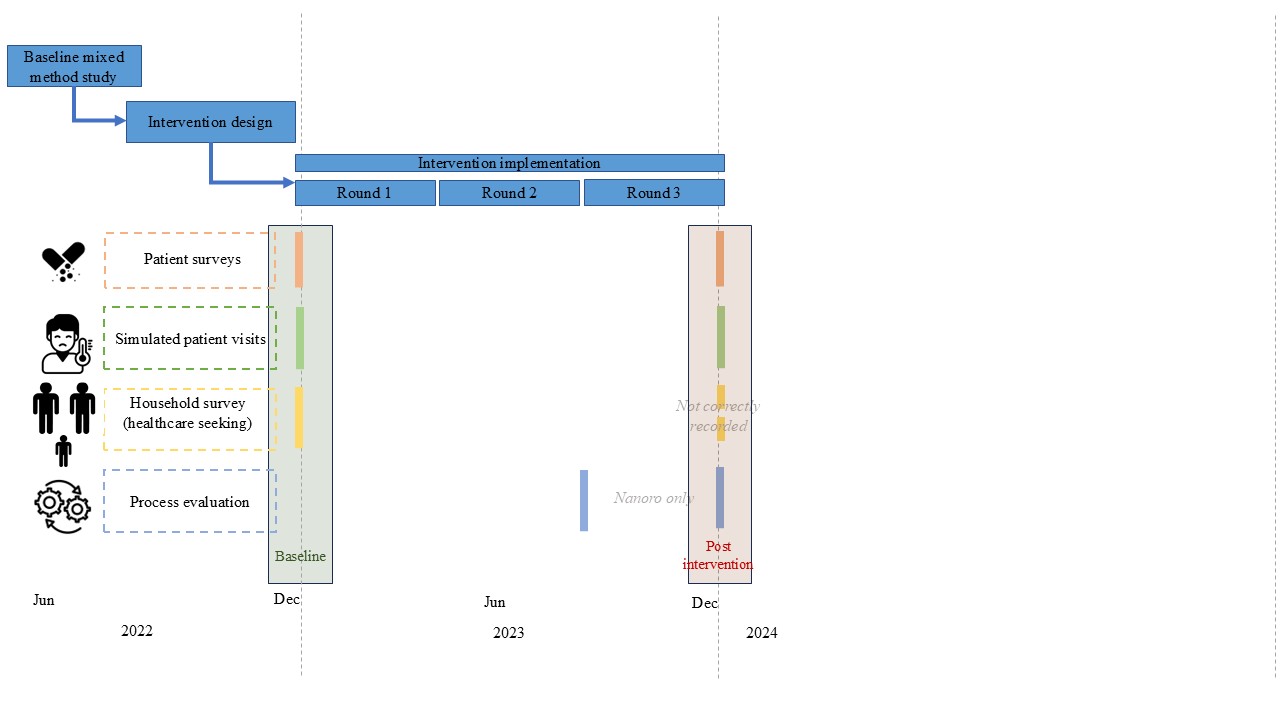

### Figure S2. Intervention and trial timeline and overview

### Supplementary text 2. Simulated patient visits scenarios and checklists to assess patient management (translated from French)

Link to original version (in French): <https://github.com/ingelbeen/cabu_intervention/blob/main/simulated_patient_visit_scenarios_checklists>

SIMULATED PATIENT VISITS clinical scenarios and checklists for assessing the quality of primary care

| 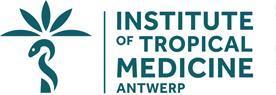 | 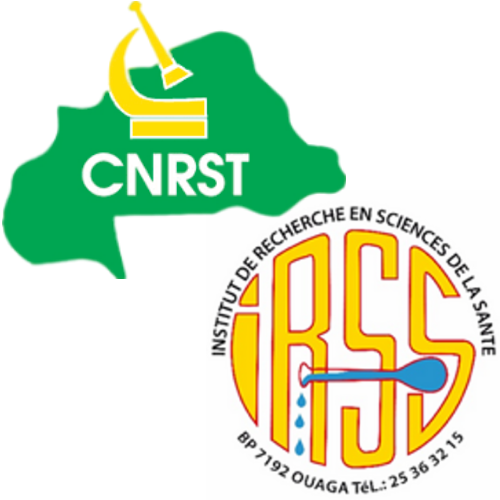 | 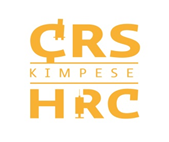 |
| --- | --- | --- |

| Study | CABU-B/C study (Optimising antibiotic use and infection control at community level through a package of behaviour-targeting interventions in Burkina Faso and DR Congo, NCT05378880) |
| --- | --- |
| Sponsor | Institute of Tropical Medicine  Nationalestraat 155  B-2000 Antwerp – Belgium |
| Authors | Brecht Ingelbeen (ITG),  Valia Daniel (CRUN),  Steven Declercq (ITG),  Marianne van der Sande (ITG), |
| Version | 1.1, 10 February 2023 (some language modifications following training of simulated patients) |

Acute gastroenteritis (non-bloody but watery). Adult presenting to the healthcare provider after a night of frequent watery stools and vomiting.

Reason for consultation: *Upon arrival*, specify only that there was watery stool during the night. Do not explicitly ask for treatment, but rather what should be done.

*Respond if asked*:

-I had 4 bowel movements during the night + 1 single episode of vomiting in the middle of the night.

-No blood in the stools (watery diarrhoea without blood), no mucus in the stools (non-mucous stools).

-No fever, no stomach ache, no cramps, no bloating, and currently no urge to vomit.

-I am feeling tired at the moment.

-There was no diarrhoea in the previous weeks.

-Generally in good health with no known chronic illnesses, HIV, and no recent malaria. No history of travel or contact with another sick person. No other cases in the household. Usually drinks water from a protected well [adapt the response according to the available and likely source of drinking water in the village]. He drank water in the morning, but did not feel abnormally thirsty.

*For this scenario, appropriate management will consist of at least (1) questions or examination of stool type or presence of blood in stool, (2) advice to stay hydrated (drink fluids, ORS), AND (3) no prescription or dispensing of antibiotics.*

|  | Yes/No/number | Max. value | Score given |
| --- | --- | --- | --- |
| **Medical history questions** *-> tick if the presence of a sign, symptom or history has been assessed* | | | |
| Type/consistency of stool |  | 2 |  |
| If stool contains blood |  | 2 |  |
| If stool contains mucus/mucous stool |  | 1 |  |
| Frequency of stools |  | 2 |  |
| Fever? History of fever? |  | 2 |  |
| Abdominal pain? Cramps? |  | 1 |  |
| Vomiting or nausea? |  | 1 |  |
| Questions about urination |  | 1 |  |
| Ability to drink, desire to drink? Thirst? |  | 1 |  |
| Lethargy and/or loss of consciousness? |  | 1 |  |
| General health condition? Immunocompromised/HIV? |  | 1 |  |
| Recent use of antibiotics? |  | 1 |  |
| Other members of the household with diarrhoea or other symptoms? |  | 1 |  |
| Source of drinking water? How is drinking water preserved at home? |  | 1 |  |
| Meal preparation? |  | 1 |  |
| Hand washing? |  | 1 |  |
| Water, sanitation and hygiene conditions in the neighbourhood or family |  | 1 |  |
| Questions about the physical environment and hygiene conditions |  | 1 |  |
| **Clinical examination** | | | |
| Takes vital signs (blood pressure, pulse or temperature) |  | 1 |  |
| Abdominal examination |  | 1 |  |
| Assessment of mucous membranes or skin pinch test (to assess for dehydration) |  | 1 |  |
| Check if the eyes are sunken |  | 1 |  |
| **Actions** | | | |
| Explain the need to stay well hydrated/drink fluids? |  | 2 |  |
| Clearly explain why it is not necessary to treat this type of acute diarrhoea with antibiotics |  | 1 |  |
| Explains the importance of safe water and food and good hygiene and sanitation practices |  | 1 |  |
| Advice on what to do if diarrhoea persists |  | 1 |  |
| **Dispensing of medicines** | | | |
| Prescribe one or more antibiotics |  | -2 |  |
| Specify the systemic antibiotics prescribed/dispensed (generic name or brand name, dose, route of administration, frequency per day, duration; exclude ointments, eye drops, or other antibiotics for external use) | | | |
| *Number of systemic antibiotics not included in the WHO 2021 essential medicines list (assessed afterwards by a clinician or pharmacist)* |  | -2* |  |
| Gives or offers to use ORS = oral rehydration solution, or zinc |  | 1 |  |
| Gives an antimalarial drug? |  | -1 |  |
| Number of drugs prescribed/dispensed for administration by injection |  | -1 |  |
| **Duration of visit** (consultation if health centre, total visit if pharmacy/vendor) in seconds |  |  |  |
| TOTAL SCORE |  |  |  |

* per Watch-group antibiotic/systemic medication given simultaneously. e.g. norfloxacin tablets = -2; ciprofloxacin p.o. and erythromycin p.o. = - 4

Acute nasopharyngitis. Adult presenting with dry cough + cold + sore throat + low-grade fever (slight feeling of warmth), all of which has been developing for 3 days but with improvement in fever on the day of the visit (history of fever).

Reason for consultation: *On arrival:* I have a cold and my body feels hot, but I took paracetamol two hours before coming to see you.

*Answer if asked:*

-The fever/hot body started yesterday (the day before the visit to the healthcare provider).

-The cough started 3 days ago.

-When I cough, I don't spit (no expectoration).

-My throat hurts and my nose is blocked.

-I have no difficulty breathing and no chest pain.

-There is also a child at home with a cough, which started four days ago (in case you are asked about other similar cases in your household).

-No other comorbidities such as HIV, hypertension, recent malaria, or diabetes (no history).

*For this scenario, appropriate management will consist of at least (1) questions or examination for the presence of respiratory signs (type and frequency of cough, sputum, difficulty breathing) AND the presence of fever; AND (2) no prescription or dispensing of antibiotics.*

|  | yes/no/number | Max. value | Score given |
| --- | --- | --- | --- |
| **Medical history questions** *-> tick if the presence of a sign, symptom or history has been assessed* | | | |
| Type of cough: dry or with sputum production |  | 1 |  |
| Duration of cough, acute onset |  | 1 |  |
| Duration of fever, or has the patient had a fever/hot body in the previous days? |  | 1 |  |
| Type of sputum (mucous? mucopurulent?) |  | 1 |  |
| Difficulty breathing or shortness of breath? |  | 1 |  |
| Sore throat? |  | 1 |  |
| Earache or ear inspection? |  | 1 |  |
| History of respiratory disease (e.g. tuberculosis, chronic obstructive pulmonary disease)? |  | 1 |  |
| **Clinical examination** | | | |
| Diagnosis of malaria (rapid diagnostic test or thick smear) |  | 1 |  |
| Inspection of throat |  | 1 |  |
| Temperature |  | 1 |  |
| Palpation of lymph nodes |  | 1 |  |
| Pulse/heart rate |  | 1 |  |
| Respiratory rate (to rule out pneumonia) |  | 1 |  |
| Auscultation |  | 1 |  |
| **Actions** | | | |
| Information that the vast majority of cases will improve after a few days/are viral in origin and do not require antibiotics |  | 1 |  |
| **Dispensing of medicines** | | | |
| Prescribes one or more antibiotics |  | - |  |
| Specify which ones (generic name or brand name, exclude ointments, eye drops, or other antibiotics for external use) | | | |
| *Number of systemic antibiotics not included in the WHO 2021 essential medicines list (assessed afterwards by a clinician or pharmacist)* |  | -2 |  |
| Prescribe an antimalarial drug? |  | -2 |  |
| Number of drugs prescribed/dispensed for injection |  | -1 |  |
| **Duration of visit** (consultation if health centre, total visit if pharmacy/vendor) in seconds |  |  |  |
| TOTAL SCORE |  |  |  |

* per Watch-group antibiotic /systemic medication given simultaneously. e.g. norfloxacin tablets = -2; ciprofloxacin p.o. and erythromycin p.o. = - 4

Severe acute pneumonia in an elderly patient: A young adult seeking care for his father, who has had a productive cough + mucous sputum + fever and headaches for 5 days.

Reason for consultation: *On arrival:* explain that he is there on behalf of his father, who feels too weak to come in due to a cough, high fever and headaches.

*Answer if asked*:

-Age: his father is 79 years old;

-Symptoms: He is currently unable to stand up and appears very tired and somewhat drowsy.

-The symptoms (cough + headache + fever) began 5 days ago. The cough, which was initially dry, became increasingly productive (with phlegm) after the second day.

- The cough worsened significantly last night, with increasingly rapid breathing that prevented him from speaking properly. Since this morning, in addition to his breathing becoming faster, he looks very tired and is drowsy.

- We have been giving him paracetamol since the second day it started.

- He does not have high blood pressure or diabetes and was fine before the cough started.

*For this scenario, proper management will consist of at least (1) questions or examination for the presence of respiratory signs (type and frequency of cough, sputum, difficulty breathing, confusion, age) AND (2) questions or examination for the presence of signs of systemic infection: presence of fever, pulse, respiratory rate AND (3) (in the case of a drop-in centre, informal vendor or CSPS) referral to a CSPS or hospital.*

|  | yes/no/number | Max. value | Score given |
| --- | --- | --- | --- |
| **Medical history questions** *-> tick if the presence of a sign, symptom or history has been assessed* | | | |
| Type of cough: dry or productive |  | 1 |  |
| Duration of cough, acute onset |  | 1 |  |
| Has the cough worsened? |  | 1 |  |
| Fever/hot body? Fever in previous days? Duration of fever |  | 1 |  |
| Sputum (mucous? mucopurulent?)? |  | 1 |  |
| Respiratory rate? Is breathing faster than usual? |  | 1 |  |
| Difficulty breathing or shortness of breath? |  | 1 |  |
| Confusion or disorientation (newly appeared)? |  | 1 |  |
| Lethargy, inability to stand up/other signs of respiratory distress? |  | 1 |  |
| History of respiratory disease (e.g. tuberculosis, chronic obstructive pulmonary disease) |  | 1 |  |
| **Actions** | | | |
| Referral to a hospital or health centre |  | 4 |  |
| Explains the need to seek appropriate care |  | 1 |  |
| **Dispensing of medicines** | | | |
| Gives (a prescription for) one or more antibiotics to treat the episode at home without transfer to hospital (not to be given during transfer to hospital or CHSC) |  | -1 |  |
| Specify the systemic antibiotics prescribed/dispensed (generic name or brand name, dose, route of administration, frequency per day, duration; exclude ointments, eye drops, or other antibiotics for external use) | | | |
| Give antimalarial medication without a malaria diagnosis (RDT/microscopy)? |  | -1 |  |
| **Duration of visit** (consultation if health centre, total visit if pharmacy/vendor) in seconds |  |  |  |
| TOTAL SCORE |  |  |  |

* per Watch-group antibiotic /systemic medication given simultaneously. e.g. norfloxacin tablets = -2; ciprofloxacin p.o. and erythromycin p.o. = - 4

Isolated acute fever: A young adult presents to the provider with fever and no other additional symptoms.

Reason for consultation: *On arrival:* I have a hot body and a headache. I had malaria three weeks ago, but the malaria treatment prescribed at the CSPS worked well.

*Answer if asked*:

-The fever started two days ago.

-The malaria I had three weeks ago was diagnosed by a rapid diagnostic test at the CSPS and treated with "A-L" (artemether/lumefantrine), which I took for three days, and I was in great shape after the treatment.

- I have no other complaints.

*For this scenario, appropriate management will consist of at least (1) questions or examination for the presence of respiratory signs; (2) a malaria test OR referral of such a patient to a CSPS or hospital or other facility with malaria diagnostic capacity; (3) if the malaria test result (negative) is available: a follow-up appointment in the following days with or without a prescription for an antipyretic.*

|  | yes/no/number | Max. value | Score given |
| --- | --- | --- | --- |
| **Medical history questions** *-> tick if the presence of a sign, symptom or history has been assessed* | | | |
| When did the signs begin? How long has the fever/high temperature lasted? |  | 1 |  |
| Abdominal complaints: diarrhoea, vomiting, stomach ache, cramps, epigastric pain? |  | 1 |  |
| Respiratory complaints: cough, nasal discharge, other respiratory complaints? |  |  |  |
| Earache? Ear discharge? |  | 1 |  |
| Sore throat? |  | 1 |  |
| Urinary/micturition pain? |  | 1 |  |
| Meningeal signs: sensitivity to light/sound, headaches? |  | 1 |  |
| Skin rash? |  | 1 |  |
| Significant impact on functioning (lethargy, etc.)? |  | 1 |  |
| History, comorbidities (HIV, hypertension, diabetes, etc.)? |  | 1 |  |
| Meals, drinking, thirst? |  | 1 |  |
| **Clinical examination** | | | |
| Temperature, pulse, heart rate, respiratory rate (at least 1) |  | 1 |  |
| Inspection of throat, with or without tongue depressor |  | 1 |  |
| Neck stiffness |  | 1 |  |
| Abdominal examination (e.g. appendicitis, etc.) |  | 1 |  |
| Blood pressure measurement |  | 1 |  |
| Auscultation |  | 1 |  |
| **Actions** | | | |
| Information that the fever may improve after a few days even without antibacterial/antiviral/antiparasitic treatment (treatment for fever is acceptable) |  | 1 |  |
| Advice to return or (go to the CSPS) if the fever worsens or other signs appear |  | 1 |  |
| Schedule a follow-up appointment or appointment for the next day for clinical follow-up |  | 1 |  |
| **Dispensing of medicines** | | | |
| Gives (a prescription for) one or more antibiotics |  | - |  |
| Specify the systemic antibiotics prescribed/dispensed (generic name or brand name, dose, route of administration, frequency per day, duration; exclude ointments, eye drops, or other antibiotics for external use) | | | |
| *Number of systemic antibiotics not included in the WHO 2021 essential medicines list (assessed afterwards by a clinician or pharmacist)* |  | -2* |  |
| Number of drugs prescribed/dispensed for injectable administration |  | -1* |  |
| Is an antimalarial drug given? |  | -1 |  |
| Specify which ones (generic name or brand name, exclude ointments, eye drops, or other antibiotics for external use) | | | |
| *This is an antimalarial drug that is not an artemisinin-based combination therapy or monotherapy with an artemisinin derivative (e.g. amodiaquine, chloroquine, etc.)* |  | -2 |  |
| **Duration of visit** (consultation if health centre, total visit if pharmacy/vendor) in seconds |  |  |  |
| TOTAL SCORE |  |  |  |

* per Watch-group antibiotic /systemic medication given simultaneously. e.g. norfloxacin tablets = -2; ciprofloxacin p.o. and erythromycin p.o. = - 4

Acute urinary tract infection: A young adult presents to the provider with burning during urination + frequent urination in small amounts (more than 7 times during the day and at least 2 times during the night).

Reason for consultation: *On arrival:* I don't feel very well and it burns when I urinate.

*Respond if asked*:

-It started yesterday morning; it burns when I urinate.

-I urinated more than seven times yesterday, but each time only a small amount came out. During the night, I also woke up twice to urinate, again only small amounts, whereas normally when I sleep at night, I don't wake up to urinate.

-I have not had unprotected sex recently.

-My water consumption has not changed either; I drink practically the same amount every day.

-This is the first time this has ever happened to me.

-Apart from this illness, I am in good health.

*For this scenario, appropriate management would consist of at least a referral to a formal health centre (CSPS or hospital) OR (1) questions about the duration and frequency of urinary complaints AND (2) treatment with antibiotics from the Access group: nitrofurantoin every 6 hours (4 doses per day) for 5 days OR smx/tmp for 3 to 8 days (8 days based on the 2008 national guideline) OR tmp for 3 days OR amoxicillin/clavulanic acid for 3 to 5 days.*

|  | yes/no/number | Max. value | Score given |
| --- | --- | --- | --- |
| **Medical history questions** *-> tick if the presence of a sign, symptom or history has been assessed* | | | |
| Duration of dysuria or burning/painful urination? When did it start? |  | 2 |  |
| Pain or discomfort in the lower abdomen? |  | 1 |  |
| Frequency of urination during the day and at night? Change in frequency? |  | 1 |  |
| Haematuria? Blood in urine? Coloured urine? |  | 1 |  |
| Presence or absence of fever/high temperature? |  | 1 |  |
| Onset/duration of fever/high temperature? |  | 1 |  |
| Vaginal discharge or irritation (to rule out a vaginal source of symptoms)? |  | 1 |  |
| Other complaints or symptoms? |  | 1 |  |
| History of urinary tract infections/painful urination? |  | 1 |  |
| History, comorbidities (HIV, hypertension, diabetes, etc.)? |  | 1 |  |
| Thirst? Drinking sufficient amounts? |  | 1 |  |
| Recent unprotected sexual contact? |  | 1 |  |
| **Clinical examination** | | | |
| Urine analysis (dipstick or microscopy) to detect bacteriuria and/or indirect signs of infection (positive leukocyte esterase and nitrites). |  | 1 |  |
| Takes temperature |  | 1 |  |
| Assess for pyelonephritis -> Specific questions about pelvic/abdominal/back/costovertebral pain |  | 1 |  |
| **Actions** | | | |
| Referral to a formal health centre (CSPS or CM) |  | 1 |  |
| Advice to return (to the CSPS) if fever worsens or other signs appear OR Schedule clinical follow-up |  | 1 |  |
| *If no antibiotics:* Information that the infection may be self-limiting  *If antibiotics are prescribed Access (see options below):* clear instructions on dosage (number of doses per day, when) and duration of treatment |  | 1 |  |
| **Dispensing of medicines** | | | |
| Specify the systemic antibiotics prescribed/dispensed (generic name or brand name, dose, route of administration, frequency per day, duration; exclude ointments, eye drops, or other antibiotics for external use) | | | |
| *Number of systemic antibiotics not included in the WHO 2021 list of essential medicines (to be assessed by a clinician or pharmacist)* |  | -2* |  |
| *Antibiotics as recommended in the WHO antibiotic book or national guidelines, with correct dosage and duration: nitrofurantoin every 6 hours (4 doses per day) for 5 days OR smx/tmp for 3 to 8 days (8 days based on the 2008 national guideline) OR tmp for 3 days OR amoxicillin/clavulanic acid for 3 to 5 days.* |  | 1 |  |
| No antibiotics |  | 1 |  |
| Number of medicines prescribed/dispensed for administration by injection |  | -1 |  |
| **Duration of visit** (consultation if health centre, total visit if pharmacy/vendor) in seconds |  |  |  |
| TOTAL SCORE |  |  |  |

* per Watch-group antibiotic /systemic medication given simultaneously. e.g. norfloxacin tablets = -2; ciprofloxacin p.o. and erythromycin p.o. = - 4

*Supplementary text 3. Methods of the process evaluation*

The process evaluation was done in five villages in Nanoro during Nov 23, 2023 – Dec 4, 2023. Participants were recruited through purposive sampling, relying on community liaisons. This recruitment method, overseen by local authorities and community leaders (village chiefs, Village Development Committees, health workers, pastors, and community informants), aimed to ensure the legitimacy and acceptability of the evaluation within the villages concerned.

A total of 12 interviews were conducted, including:

- 2 focus group discussions with men and women from the community,
- 10 in-depth individual interviews with key stakeholders, including religious leaders, community leaders, and informal medicine vendors.

Combining focus group discussions and individual interviews allowed for the integration of community perspectives with those of influential stakeholders. Data were collected in the local language, then transcribed into French and translated into English. The analysis was carried out manually, using a thematic approach aimed at highlighting the perceptions, practices and social dynamics related to the intervention.

### Supplementary text 4. Estimation of statistical power to observe a reduction in Watch-group antibiotic use.

To ensure differences in the prevalence of Watch-group antibiotic use at baseline between intervention and control clusters are factored in the analysis, the planned analysis was a difference-in-differences (net difference) analysis, comparing interventions vs control while adjusting for baseline, to reduce residual variance and the required sample size. We explored the effect of different scenarios, i.e. different values of the assumed effect size (reduction in Watch-group antibiotics by 40%), of the baseline prevalence of Watch-group antibiotic use (24%), and of intra class correlation (ICC), on statistical power to observe the difference in prevalence of Watch-group antibiotic use. We assumed 11 intervention vs control clusters, allowing a site-specific analysis. The script with the estimation of statistical power can be found here: <https://github.com/esthervankleef/sample_size_jpiamr/blob/master/Scripts/main_trial_binary.R>

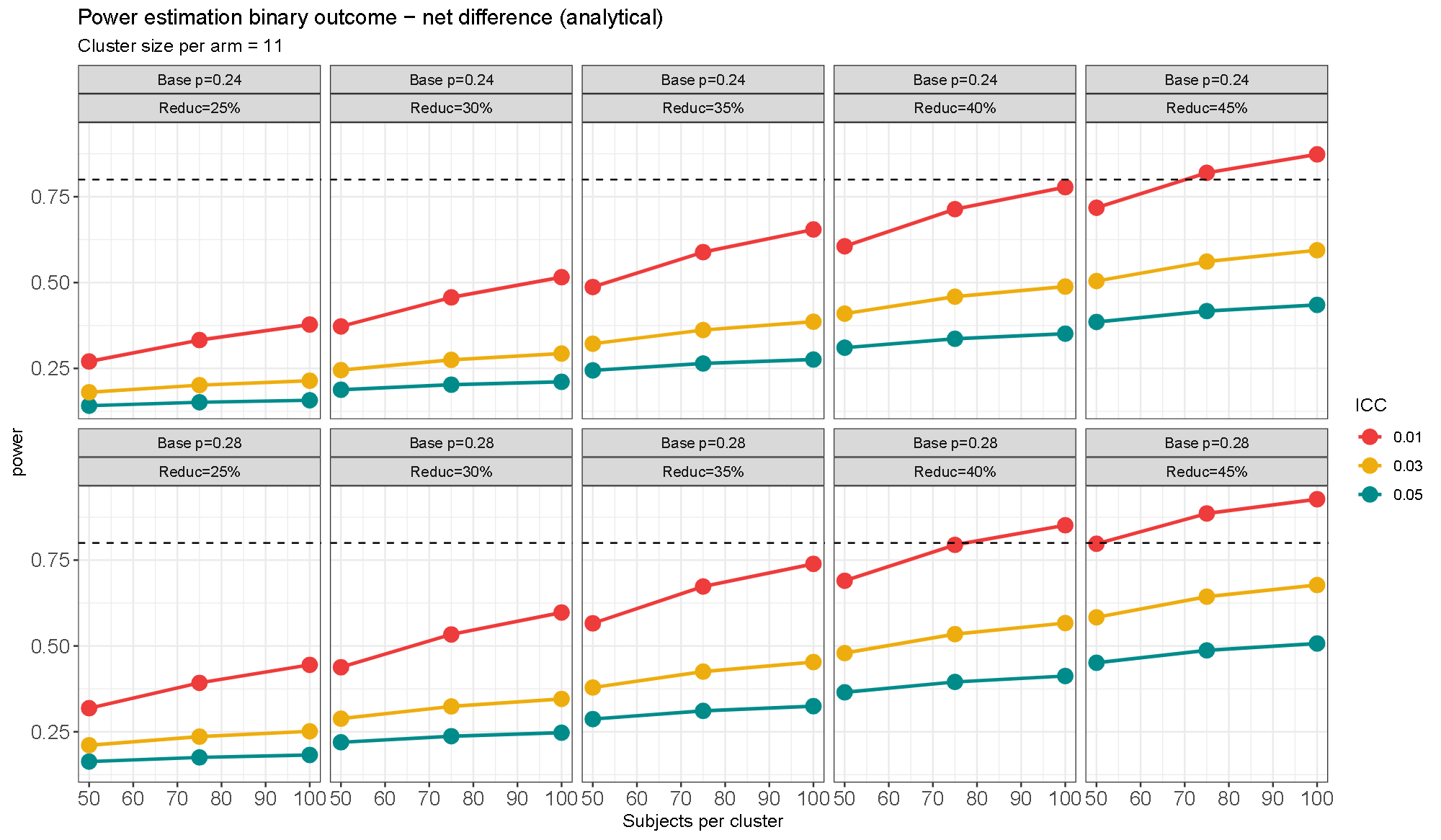

### Table S1. Community-level healthcare visits per 1000 inhabitants per month estimated from household survey data. Jan 24, 2023 to Apr 6, 2023 (Kimpese) and Oct 11, 2022 to Feb 4, 2023 (Nanoro).

| Type of provider | Reported visits in the last 3 months during prior survey* | | | Reported visits in the last 3 months | | | Reported visits in the last month | | |
| --- | --- | --- | --- | --- | --- | --- | --- | --- | --- |
|  | n | rate | 95%CI | n | rate | 95%CI | n | rate | 95%CI |
| **Kimpese (DR Congo)** | N=31221 person-months | | |  | | | N=1326 person-months | | |
| Health centre |  | 25·5 | 24·6-26·4 |  |  |  | 83 | 62·6 | 49·1-76·1 |
| Private clinic£ |  | 31·0 | 30·0-32·0 |  |  |  |  |  |  |
| Community pharmacy/store |  | 17·6 | 16·9-18·3 |  |  |  | 35 | 26·4 | 17·7-35·1 |
| Informal vendor |  | § |  |  |  |  | § | - | - |
| Traditional healer |  | 13·4 | 12·8-14·0 |  |  |  | 1 | 0·8 | -0·7-2·2 |
| Hospital# |  | 1·2 | 1·0-1·4 |  |  |  |  |  |  |
| OVERALL |  | 88·7 | 81·9-95·4 |  |  |  | 119 | 89·7 | 73·6-105 |
| **Nanoro (Burkina Faso)** | N=32079 person-months | | | N=11166 person-months | | | N=3722 person-months | | |
| Health centre | 1328 | 41·4 |  | 232 | 20·8 | 18·1-23·5 | 98 | 26·3 | 21·1-31·5 |
| Community pharmacy/store | 89 | 2·8 |  | 28 | 2·5 | 1·6-3·4 | 9 | 2·4 | 0·8-4·0 |
| Informal vendor | 315 | 9·8 |  | 54 | 4·8 | 3·5-6·1 | 20 | 5·4 | 3·0-7·7 |
| Traditional healer | 62 | 1·9 |  | 4 | 0·4 | 0·0-0·7 | 2 | 0·5 | -0·2-1·3 |
| OVERALL | 1794 | 56·0 |  | 318 | 28·5 | 19·1-37·9 | 129 | 34·7 | 28·7-40·6 |

*The prior surveys were limited to four villages in DRC (rural Viaza, Malanga, Kavuaya, and peri-urban Nkandu) during November 2019-June 2020, and two in Burkina Faso (rural Nazoanga and peri-urban Nanoro) during October 2020-December 2021. Results were published [doi.org/10.1016/j.cmi.2022.04.002](http://doi.org/10.1016/j.cmi.2022.04.002) and [doi.org/10.1007/978-981-16-3787-2_4](http://doi.org/10.1007/978-981-16-3787-2_4)

£During the 2022 household survey in Kimpese, it was not specified whether primary care clinics were public health centres or private clinics, and the specific moment of healthcare seeking during the past month could not be reliably recorded.

§In Kimpese, no informal vendors were identified and community pharmacies or stores often lack qualified staff.

#Healthcare visits to secondary care were only recorded in the prior healthcare utilisation surveys.

### Table S2. Characteristics of participants to patient surveys, by site, round, and intervention group, Oct 2022-Feb 2023 and Oct 2023-Mar 2024

| **Patient characteristic** | | **Kimpese** | | | | | | | | **Nanoro** | | | | | | | |
| --- | --- | --- | --- | --- | --- | --- | --- | --- | --- | --- | --- | --- | --- | --- | --- | --- | --- |
|  |  | **baseline** | | | | **endline** | | | | **baseline** | | | | **endline** | | | |
|  |  | **control** | | **intervention** | | **control** | | **intervention** | | **control** | | **intervention** | | **control** | | **intervention** | |
|  |  | **n** | **%** | **n** | **%** | **n** | **%** | **n** | **%** | **n** | **%** | **n** | **%** | **n** | **%** | **n** | **%** |
| Type of visit | Acute illness | 626 | 96·2 | 741 | 94·4 | 886 | 93·6 | 691 | 89·6 | 1216 | 80·7 | 1794 | 87·5 | 1246 | 90·5 | 1647 | 91·3 |
|  | Chronic illness | 14 | 2·1 | 15 | 1·9 | 40 | 4·2 | 67 | 8·7 | 123 | 8·2 | 101 | 4·9 | 30 | 2·2 | 43 | 2·4 |
|  | No illness (e.g. refill) | 11 | 1·7 | 29 | 3·7 | 21 | 2·2 | 13 | 1·7 | 45 | 3·0 | 34 | 1·7 | 12 | 0·9 | 11 | 0·6 |
|  | Animal health |  |  |  |  |  |  |  |  | 123 | 8·2 | 122 | 5·9 | 89 | 6·5 | 102 | 5·7 |
| Age group* | 0-4 years | 138 | 22·0 | 182 | 24·6 | 184 | 20·8 | 158 | 22·9 | 112 | 9·2 | 216 | 12·0 | 154 | 12·4 | 270 | 16·4 |
|  | 5-17 years | 128 | 20·4 | 168 | 22·7 | 192 | 21·7 | 162 | 23·4 | 101 | 8·3 | 179 | 10·0 | 139 | 11·2 | 208 | 12·6 |
|  | 18-64 years | 337 | 53·8 | 365 | 49·3 | 496 | 56·0 | 354 | 51·2 | 994 | 81·7 | 1369 | 76·3 | 927 | 74·4 | 1116 | 67·8 |
|  | >/= 65 years | 23 | 3·7 | 26 | 3·5 | 14 | 1·6 | 17 | 2·5 | 9 | 0·7 | 30 | 1·7 | 26 | 2·1 | 53 | 3·2 |
| Sex* | Female | 324 | 51·8 | 400 | 54·0 | 452 | 51·0 | 352 | 50·9 | 611 | 50·2 | 879 | 49·0 | 537 | 43·0 | 732 | 44·0 |
|  | Male | 302 | 48·2 | 341 | 46·0 | 434 | 49·0 | 339 | 49·1 | 605 | 49·8 | 915 | 51·0 | 709 | 57·0 | 915 | 56·0 |
| Assigned infection* | Malaria | 195 | 31·1 | 256 | 34·5 | 287 | 32·4 | 316 | 45·7 | 164 | 13·5 | 346 | 19·3 | 172 | 13·8 | 375 | 22·8 |
|  | Acute respiratory infection  (other than pneumonia) | 88 | 14·1 | 71 | 9·6 | 78 | 8·8 | 58 | 8·4 | 418 | 34·4 | 592 | 33·0 | 392 | 31·5 | 438 | 26·6 |
|  | Skin/soft tissue infection | 45 | 7·2 | 50 | 6·8 | 81 | 9·1 | 51 | 7·4 | 1 | 0·1 | 17 | 0·9 | 6 | 0·5 | 11 | 0·7 |
|  | Unexplained fever | 53 | 8·5 | 83 | 11·2 | 63 | 7·1 | 43 | 6·2 | 161 | 13·2 | 213 | 11·9 | 198 | 15·9 | 217 | 13·2 |
|  | Unexplained gastro-intestinal complaints | 40 | 6·4 | 50 | 6·8 | 34 | 3·8 | 19 | 2·8 | 47 | 3·9 | 67 | 3·7 | 55 | 4·4 | 60 | 3·6 |
|  | Non-bacterial infectious (viral outbreak, worms, amoebae) | 32 | 5·1 | 34 | 4·6 | 51 | 5·8 | 25 | 3·6 | 2 | 0·2 | 1 | 0·1 |  |  |  |  |
|  | Other non-specific symptoms | 20 | 3·2 | 40 | 5·4 | 96 | 10·8 | 48 | 7·0 | 289 | 23·8 | 341 | 19·0 | 251 | 20·1 | 302 | 18·3 |
|  | Typhoid fever or sepsis | 35 | 5·6 | 29 | 3·9 | 58 | 6·6 | 34 | 4·9 | 1 | 0·1 | 6 | 0·3 |  |  | 1 | 0·1 |
|  | Urinary tract infection | 9 | 1·4 | 20 | 2·7 | 27 | 3·0 | 15 | 2·2 | 35 | 2·9 | 21 | 1·2 | 62 | 5·0 | 79 | 4·8 |
|  | Gastroenteritis | 18 | 2·9 | 19 | 2·6 | 16 | 1·8 | 15 | 2·2 | 29 | 2·4 | 79 | 4·4 | 61 | 4·9 | 68 | 4·1 |
|  | Pneumonia | 2 | 0·3 | 1 | 0·1 | 34 | 3·8 | 26 | 3·8 | 1 | 0·1 | 1 | 0·1 |  |  |  |  |
|  | Sexually transmitted infection | 4 | 0·6 | 6 | 0·8 | 3 | 0·3 | 2 | 0·3 |  |  |  |  |  |  |  |  |
|  | Dental  infections | 6 | 1·0 |  |  | 2 | 0·2 | 1 | 0·1 | 7 | 0·6 | 6 | 0·3 | 1 | 0·1 | 2 | 0·1 |
|  | Other | 73 | 11·7 | 71 | 9·6 | 48 | 5·4 | 35 | 5·1 | 61 | 5·0 | 104 | 5·8 | 48 | 3·9 | 94 | 5·7 |
|  | Not recorded | 6 | 1·0 | 11 | 1·5 | 8 | 0·9 | 3 | 0·4 |  |  |  |  |  |  |  |  |

*among patients visiting with acute illness

### Table S3. Frequency, crude and cluster- and provider-weighted prevalence of (Watch-group) antibiotic use, overall, by site and by type of provider

| **Subgroup** | **control** | | | | | | | | **intervention** | | | | | | | | | | **Adjusted risk ratio** | | |
| --- | --- | --- | --- | --- | --- | --- | --- | --- | --- | --- | --- | --- | --- | --- | --- | --- | --- | --- | --- | --- | --- |
|  | **baseline** | | | | **endline** | | | | **baseline** | | | | | **endline** | | | | |  |  |  |
|  | **n** | **crude %** | **weighted %** | **95%CI** | **n** | **crude %** | **weighted %** | **95%CI** | **n** | | **crude %** | **weighted %** | **95%CI** | | **n** | **crude %** | **weighed %** | **95%CI** | | **aRR** | **95%CI** |
| **ANY ANTIBIOTIC USE** | | | | | | | | | | | | | | | | | | | | | |
| **Overall**, weighted | 711/2092 | 34·0 | 39·7 | 26·1-53·2 | 680/2132 | 31·9 | 42·0 | 25·7-58·4 | | 1107/2787 | 39·7 | 56·2 | 35·9-76·5 | | 617/2338 | 26·4 | 37·5 | 28·3 46·7 | | 0·48 | 0·28-0·82 |
| **By site** |  |  |  |  |  |  |  |  | |  |  |  |  | |  |  |  |  | |  |  |
| Kimpese | 495/876 | 56·5 | 51·9 | 32·3-71·5 | 424/886 | 47·9 | 50·3 | 30·0-70·7 | | 671/993 | 67·6 | 73·5 | 55·2-91·8 | | 309/691 | 44·7 | 49·3 | 39·4 59·1 | | 0·53 | 0·28-0·99 |
| Nanoro | 216/1216 | 17·8 | 21·4 | 16·7-26·2 | 256/1246 | 20·5 | 19·3 | 10·5-28 | | 436/1794 | 24·3 | 28·5 | 22·1-34·9 | | 308/1647 | 18·7 | 22·2 | 15·1 29·3 | | 0·40 | 0·20-0·79 |
| **By type of provider** |  |  |  |  |  |  |  |  | |  |  |  |  | |  |  |  |  | |  |  |
| health centre | 246/744 | 33·1 | 33·2 | 23·8-42·6 | 203/664 | 30·6 | 28·6 | 13·7-43·5 | | 484/1224 | 39·5 | 41·0 | 29·0-53·0 | | 247/1035 | 23·9 | 25·9 | 17·9 33·9 | | 0·47 | 0·24-0·91 |
| private clinic | 109/139 | 78·4 | 78·4 | 78·4-78·4 | 199/273 | 72·9 | 74·4 | 71·0-77·8 | | 143/211 | 67·8 | 87·3 | 77·0-97·7 | | 97/178 | 54·5 | 52·8 | 35·9 69·8 | | 0·43 | 0·14-1·3 |
| private pharmacy | 296/592 | 50 | 46·8 | 17·6-75·9 | 220/602 | 36·5 | 26·1 | 10·4-41·8 | | 355/565 | 62·8 | 51·1 | 30·5-71·6 | | 166/393 | 42·2 | 45·5 | 38·8 52·2 | | 0·78 | 0·30-2·0 |
| Informal vendor | 60/617 | 9·7 | 9·1 | 5·1-13·1 | 58/593 | 9·8 | 9·2 | 5·8-12·6 | | 125/787 | 15·9 | 16·4 | 10·7-22·0 | | 107/732 | 14·6 | 15·2 | 10·1 20·3 | | 0·81 | 0·49-1·4 |
| **WATCH-GROUP ANTIBIOTIC USE** | | | | | | | | | | | | | | | | | | | | | |
| **Overall**, weighted | 211/2092 | 10·1 | 13·4 | 4·8-22·0 | 197/2132 | 9·2 | 21·2 | 8·9-33·5 | | 389/2787 | 14·0 | 26·8 | 8·8-44·8 | | 189/2338 | 8·1 | 17·1 | 7·7-26·5 | | 0·33 | 0·14-0·78 |
| **By site** |  |  |  |  |  |  |  |  | |  |  |  |  | |  |  |  |  | |  |  |
| Kimpese | 182/876 | 20·8 | 20·0 | 7·5-32·6 | 170/886 | 19·2 | 28·3 | 13·9-42·7 | | 347/993 | 34·9 | 41·6 | 24·7-58·4 | | 165/691 | 23·9 | 28·9 | 18·5-39·4 | | 0·34 | 0·18-0·67 |
| Nanoro | 29/1216 | 2·4 | 3·5 | 1·6-5·4 | 27/1246 | 2·2 | 1·6 | 0·5-2·8 | | 42/1794 | 2·3 | 3·3 | 1·3-5·2 | | 24/1647 | 1·5 | 1·7 | 0·6-2·8 | | 0·27 | 0·05-1·34 |
| **By type of provider** |  |  |  |  |  |  |  |  | |  |  |  |  | |  |  |  |  | |  |  |
| health centre | 62/744 | 8·3 | 9·4 | 1·8-17·1 | 26/664 | 3·9 | 3·7 | 0·6-6·9 | | 144/1224 | 11·8 | 11·9 | 2·6-21·2 | | 47/1035 | 4·5 | 4·2 | 0·5-7·8 | | 0·50 | 0·21-1·18 |
| private clinic | 70/139 | 50·4 | 50·4 | 50·4-50·4 | 105/273 | 38·5 | 47·8 | 38·4-57·1 | | 96/211 | 45·5 | 56·5 | 50·7-62·4 | | 69/178 | 38·8 | 38·1 | 24·5-51·7 | | 0·68 | 0·28-1·69 |
| private pharmacy | 76/592 | 12·8 | 12·5 | 1·4-23·6 | 66/602 | 11·0 | 11·3 | 5·2-17·4 | | 141/565 | 25·0 | 17·4 | 6·7-28·1 | | 70/393 | 17·8 | 18·0 | 10·4-25·5 | | 0·40 | 0·14-1·16 |
| Informal vendor | 3/617 | 0·5 | 0·5 | 0-1·3 | 0/593 | 0 | 0 | 0-0 | | 8/787 | 1·0 | 0·9 | 0-2·0 | | 3/732 | 0·4 | 0·4 | 0-0·8 | | 1·1* | 0·41-2·8 |

* because there is a subgroup with 0 patients with Watch-group antibiotic use, we added a small continuity correction

### Table S4. Frequency, crude and cluster- and provider-weighted prevalence of (Watch-group) antibiotic use by assigned infection

| **Assigned infection** | **control** | | | | | | | | **intervention** | | | | | | | | | **Adjusted risk ratio** | | |
| --- | --- | --- | --- | --- | --- | --- | --- | --- | --- | --- | --- | --- | --- | --- | --- | --- | --- | --- | --- | --- |
|  | **baseline** | | | | **endline** | | | | **baseline** | | | | **endline** | | | | |  |  |  |
|  | **n** | **crude %** | **weighed %** | **95%CI** | **n** | **crude %** | **weighed %** | **95%CI** | **n** | **crude %** | **weighed %** | **95%CI** | | **n** | **crude %** | **weighed %** | **95%CI** | | **aRR** | **95%CI** |
| **ANY ANTIBIOTIC USE** |  |  |  |  |  |  |  |  |  |  |  |  | |  |  |  |  | |  |  |
| malaria | 118/381 | 31·0 | 39·3 | 20·2-58·3 | 151/459 | 32·9 | 38·9 | 26·2-51·5 | 238/639 | 37·2 | 44·8 | 29·9-59·7 | | 115/691 | 16·6 | 22·1 | 12·3-31·9 | | 0·29 | 0·13-0·65 |
| acute respiratory infection | 135/535 | 25·2 | 35·2 | 27·5-43·0 | 128/470 | 27·2 | 36·4 | 23·6-49·1 | 182/684 | 26·6 | 38·4 | 27·1-49·6 | | 115/496 | 23·2 | 30·6 | 21·5-39·7 | | 0·70 | 0·32-1·5 |
| skin/soft tissue infection | 71/84 | 84·5 | 89·1 | 79·3-98·9 | 59/87 | 67·8 | 67·7 | 53·3-82·1 | 85/97 | 87·6 | 89·8 | 83·1-96·5 | | 46/62 | 74·2 | 79·6 | 66·1-93·0 | | 0·96 | 0·41-2·3 |
| unexplained fever | 68/252 | 27·0 | 35·2 | 18·8-51·6 | 41/261 | 15·7 | 24·3 | 9·9-38·6 | 111/324 | 34·3 | 50·7 | 31·8-69·6 | | 41/260 | 15·8 | 30·7 | 18·2-43·1 | | 0·36 | 0·14-0·92 |
| unexplained gastro-intestinal | 25/103 | 24·3 | 28·3 | 12·8-43·8 | 14/89 | 15·7 | 24·5 | 8·4-40·5 | 48/131 | 36·6 | 55·6 | 40·0-71·2 | | 13/79 | 16·5 | 22·8 | 4·1-41·5 | | 0·59 | 0·15-2·3 |
| non-bacterial infectious (viral outbreak, scabies, worms, amoebae) | 33/49 | 67·3 | 68·8 | 60·7-76·8 | 26/51 | 51·0 | 60·1 | 33·4-86·9 | 51/63 | 81·0 | 81·8 | 72·6-91·1 | | 19/25 | 76·0 | 75·1 | 52·7-97·6 | | 0·41 | 0·11-1·6 |
| non-specific symptoms or complaints | 48/313 | 15·3 | 26·1 | 18·0-34·1 | 37/347 | 10·7 | 15·0 | 4·2-25·7 | 79/390 | 20·3 | 38·9 | 24·4-53·3 | | 44/350 | 12·6 | 20·1 | 10·4-29·8 | | 0·95 | 0·38-2·4 |
| typhoid or sepsis | 48/49 | 98·0 | 99·9 | 99·5-100 | 57/58 | 98·3 | 98·1 | 94·9-100 | 58/58 | 100·0 | 100·0 | 100-100 | | 33/35 | 94·3 | 95·8 | 89·9-100 | | 0·37 | 0·12-1·1 |
| urinary tract infection | 32/67 | 47·8 | 80·2 | 61·3-99·0 | 30/89 | 33·7 | 64·5 | 39·1-89·9 | 34/56 | 60·7 | 87·1 | 79·5-94·6 | | 15/94 | 16·0 | 51·7 | 21·5-81·9 | | 0·54 | 0·16-1·9 |
| gastroenteritis | 34/59 | 57·6 | 75·6 | 57·3-93·9 | 47/77 | 61·0 | 65·0 | 47·8-82·2 | 72/109 | 66·1 | 78·2 | 66·7-89·6 | | 54/83 | 65·1 | 76·0 | 60·0-92·1 | | 0·38 | 0·13-1·1 |
| pneumonia | 3/4 | 75·0 | 76·5 | 34·6-100 | 27/34 | 79·4 | 82·4 | 63·1-100 | 6/7 | 85·7 | 85·8 | 57·7-100 | | 25/26 | 96·2 | 96·4 | 90·3-100 | | 1·57 | 0·55-4·5 |
| sexually transmitted infection | 8/8 | 100·0 | 100·0 | 100-100 | 2/3 | 66·7 | 49·6 |  | 15/16 | 93·8 | 86·8 | 60·8-100 | | 1/2 | 50·0 | 37·4 |  | | 0·11 | 0·01-0·43 |
| dental | 5/14 | 35·7 | 77·7 | 46·8-100 | 2/3 | 66·7 | 68·8 | 16·9-100 | 1/7 | 14·3 | 40·5 | 0-94·1 | | 2/3 | 66·7 | 37·5 | 0-94·2 | | 2·93 | 0·84-10 |
| other | 74/165 | 44·8 | 44·2 | 29·7-58·7 | 54/96 | 56·2 | 48·5 | 33·6-63·5 | 114/191 | 59·7 | 54·1 | 46·3-61·8 | | 92/129 | 71·3 | 49·5 | 36·3-62·8 | | 1·50 | 0·77-2·9 |
| NA | 9/9 | 100·0 |  |  | 5/8 | 62·5 |  |  | 13/15 | 86·7 |  |  | | 2/3 | 66·7 |  |  | |  |  |
| **WATCH-GROUP ANTIBIOTIC USE** | |  |  |  |  |  |  |  |  |  |  |  | |  |  |  |  | |  |  |
| malaria | 118/381 | 31·0 | 39·3 | 20·2-58·3 | 151/459 | 32·9 | 38·9 | 26·2-51·5 | 238/639 | 37·2 | 44·8 | 29·9-59·7 | | 115/691 | 16·6 | 22·1 | 12·3-31·9 | | 0·54 | 0·19-1·6 |
| acute respiratory infection | 135/535 | 25·2 | 35·2 | 27·0·5-43 | 128/470 | 27·2 | 36·4 | 23·6-49·1 | 182/684 | 26·6 | 38·4 | 27·1-49·6 | | 115/496 | 23·2 | 30·6 | 21·5-39·7 | | 0·23 | 0·05-1·1 |
| skin/soft tissue infection | 71/84 | 84·5 | 89·1 | 79·3-98·9 | 59/87 | 67·8 | 67·7 | 53·3-82·1 | 85/97 | 87·6 | 89·8 | 83·1-96·5 | | 46/62 | 74·2 | 79·6 | 66·1-93·0 | | 0·48 | 0·18-1·3 |
| unexplained fever | 68/252 | 27·0 | 35·2 | 18·8-51·6 | 41/261 | 15·7 | 24·3 | 9·9-38·6 | 111/324 | 34·3 | 50·7 | 31·8-69·6 | | 41/260 | 15·8 | 30·7 | 18·2-43·1 | | 0·29 | 0·10-0·87 |
| unexplained gastro-intestinal | 25/103 | 24·3 | 28·3 | 12·8-43·8 | 14/89 | 15·7 | 24·5 | 8·4-40·5 | 48/131 | 36·6 | 55·6 | 40-71·2 | | 13/79 | 16·5 | 22·8 | 4·1-41·5 | | 0·21 | 0·02-1·9 |
| non-bacterial infectious (viral outbreak, scabies, worms, amoebae) | 33/49 | 67·3 | 68·8 | 60·7-76·8 | 26/51 | 51·0 | 60·1 | 33·4-86·9 | 51/63 | 81·0 | 81·8 | 72·6-91·1 | | 19/25 | 76·0 | 75·1 | 52·7-97·6 | | 1·3 | 0·36-4·9 |
| non-specific symptoms or complaints | 48/313 | 15·3 | 26·1 | 18·0-34·1 | 37/347 | 10·7 | 15·0 | 4·2-25·7 | 79/390 | 20·3 | 38·9 | 24·4-53·3 | | 44/350 | 12·6 | 20·1 | 10·4-29·8 | | 1·2 | 0·22-6·8 |
| typhoid or sepsis | 48/49 | 98·0 | 99·9 | 99·5-100 | 57/58 | 98·3 | 98·1 | 94·9-100 | 58/58 | 100·0 | 100·0 | 100-100 | | 33/35 | 94·3 | 95·8 | 89·9-100 | | 0·33 | 0·09-1·2 |
| urinary tract infection | 32/67 | 47·8 | 80·2 | 61·3-99·0 | 30/89 | 33·7 | 64·5 | 39·1-89·9 | 34/56 | 60·7 | 87·1 | 79·5-94·6 | | 15/94 | 16·0 | 51·7 | 21·5-81·9 | | 0·19 | 0·04-0·95 |
| gastroenteritis | 34/59 | 57·6 | 75·6 | 57·3-93·9 | 47/77 | 61·0 | 65·0 | 47·8-82·2 | 72/109 | 66·1 | 78·2 | 66·7-89·6 | | 54/83 | 65·1 | 76·0 | 60·0-92·1 | | 0·75 | 0·19-3·0 |
| pneumonia | 3/4 | 75·0 | 76·5 | 34·6-100 | 27/34 | 79·4 | 82·4 | 63·1-100 | 6/7 | 85·7 | 85·8 | 57·7-100 | | 25/26 | 96·2 | 96·4 | 90·3-100 | |  |  |
| sexually transmitted infection | 8/8 | 100·0 | 100·0 | 100-100 | 2/3 | 66·7 | 49·6 | 0-100 | 15/16 | 93·8 | 86·8 | 60·8-100 | | 1/2 | 50·0 | 37·4 | 0-100 | | 0·13 | 0·02-0·81 |
| dental | 5/14 | 35·7 | 77·7 | 46·8-100 | 2/3 | 66·7 | 68·8 | 16·9-100 | 1/7 | 14·3 | 40·5 | 0-94·1 | | 2/3 | 66·7 | 37·5 | 0-94·2 | |  |  |
| other | 74/165 | 44·8 | 44·2 | 29·7-58·7 | 54/96 | 56·2 | 48·5 | 33·6-63·5 | 114/191 | 59·7 | 54·1 | 46·3-61·8 | | 92/129 | 71·3 | 49·5 | 36·3-62·8 | | 1·2 | 0·41-3·5 |
| NA | 9/9 | 100·0 |  |  | 5/8 | 62·5 |  |  | 13/15 | 86·7 |  |  | | 2/3 | 66·7 |  |  | |  |  |

### Table S5. Rate of antibiotic use episodes per 1000 inhabitants per month, by site and by type of healthcare provider, before and after intervention in intervention clusters.

|  |  |  | **Any antibiotic use** | | **Watch-group antibiotic use** | |
| --- | --- | --- | --- | --- | --- | --- |
| **site** | **provider type** | **round** | **rate** | **95% CI** | **rate** | **95% CI** |
| Kimpese | health centre | baseline | 17·8 | 13·9 - 22·8 | 8·9 | 6·7 - 11·8 |
|  |  | post | 9·8 | 6·2 - 15·6 | 4·9 | 1·6 - 14·6 |
|  | private clinic | baseline | 30·0 | 23·4 - 38·5 | 19·4 | 15·2 - 24·7 |
|  |  | post | 18·1 | 12·2 - 27·0 | 13·1 | 8·5 - 20·2 |
|  | private pharmacy | baseline | 14·1 | 8·1 - 24·7 | 4·9 | 2·2 - 10·9 |
|  |  | post | 12·7 | 8·9 - 18·2 | 5·1 | 3·0 - 8·7 |
| Nanoro | health centre | baseline | 8·4 | 6·2 - 11·5 | 1·0 | 0·5 - 2·1 |
|  |  | post | 6·4 | 4·1 - 9·9 | 0·5 | 0·2 - 1·5 |
|  | informal vendor | baseline | 0·9 | 0·5 - 1·6 | 0·0 | 0·0 - NaN |
|  |  | post | 0·8 | 0·4 - 1·4 | 0·0 | 0·0 - NaN |
|  | private pharmacy | baseline | 0·6 | 0·3 - 1·4 | 0·1 | 0·0 - 0·3 |
|  |  | post | 0·4 | 0·2 - 0·9 | 0·1 | 0·0 - 0·5 |

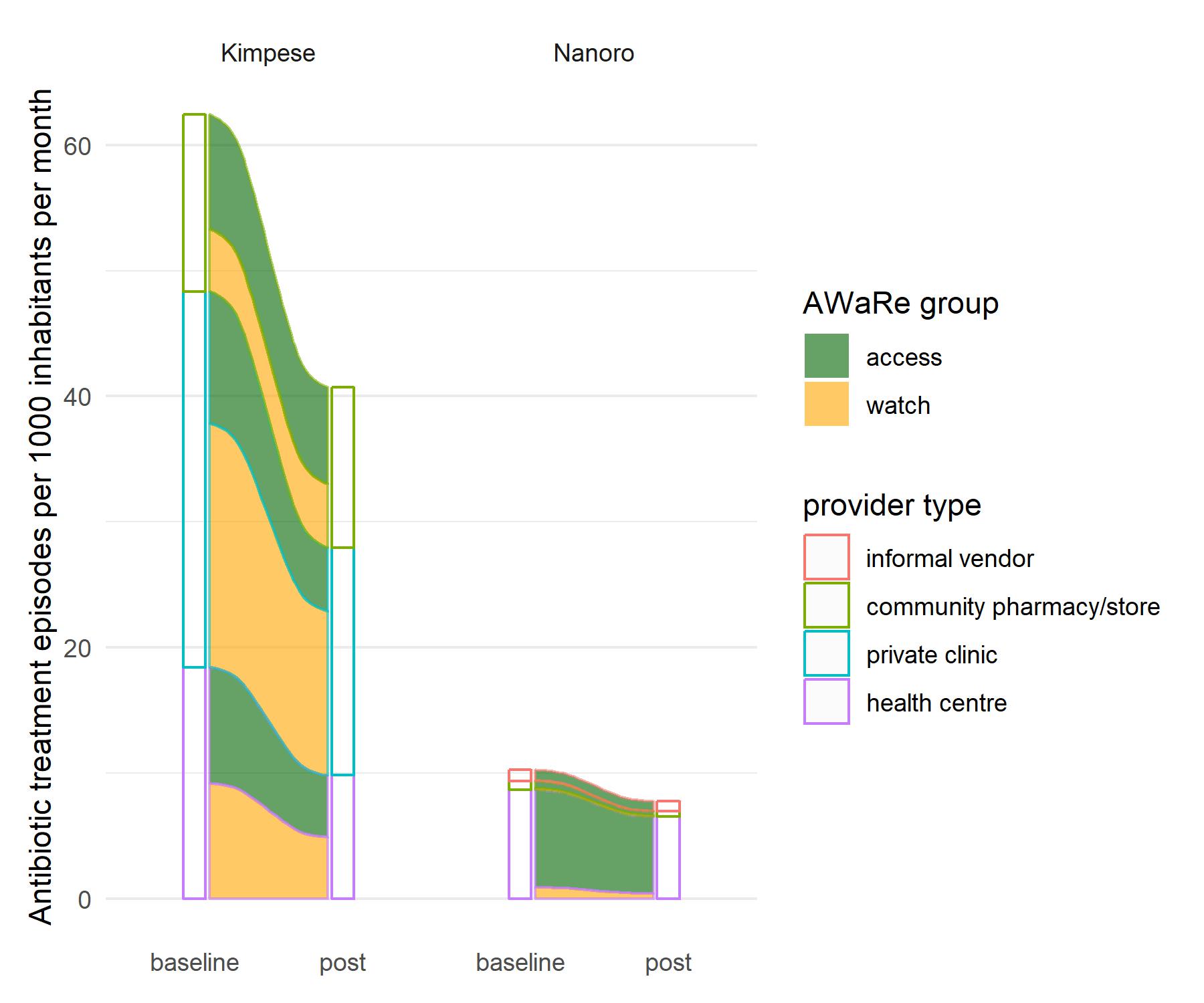

### Figure S3. Rate of antibiotic use episodes per 1000 inhabitants per month, before and after the intervention bundle in intervention clusters.

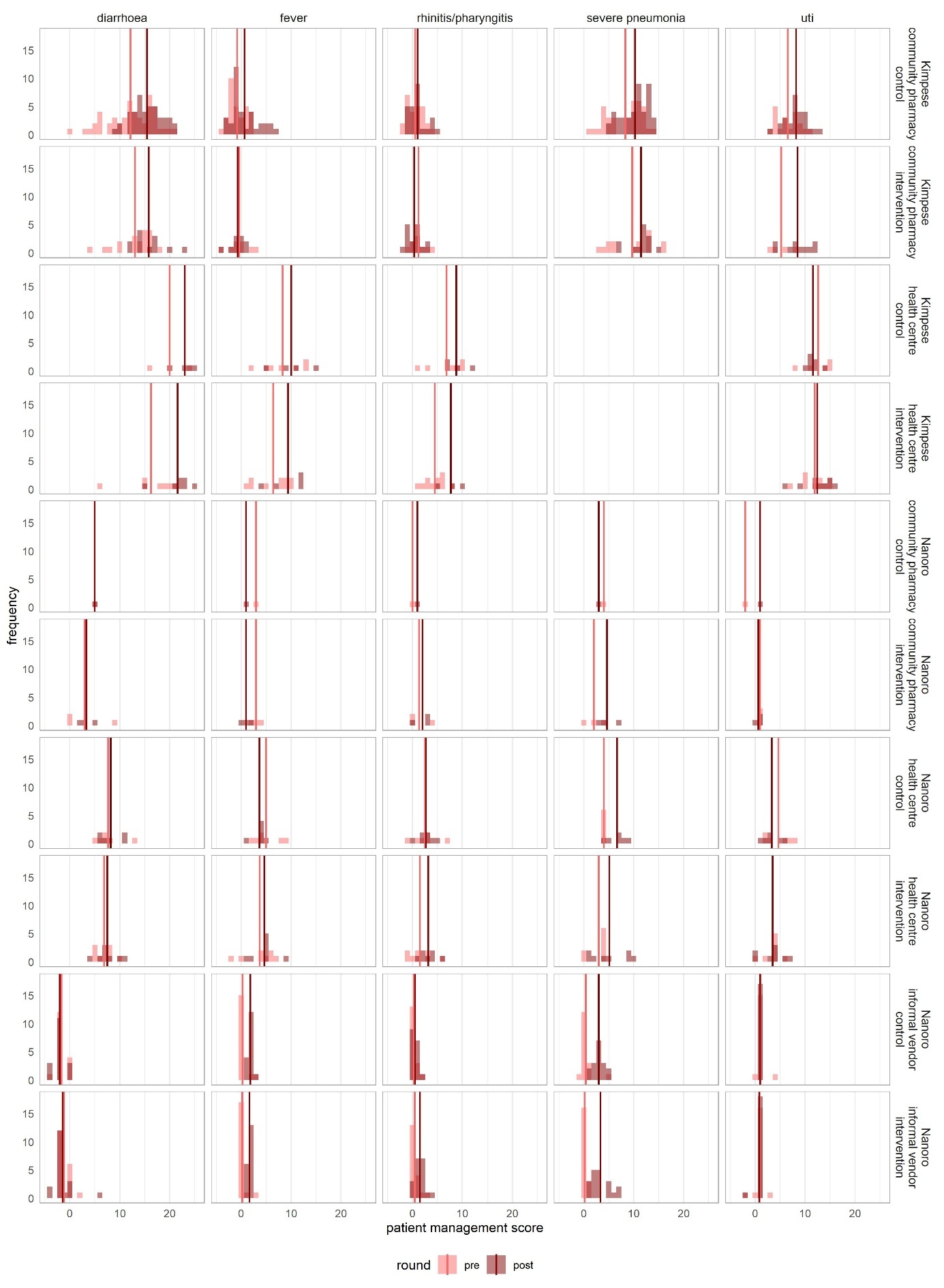

### Figure S4. Patient management scores at all healthcare providers in the intervention group before vs after intervention, faceted by site, by type of provider, by intervention/control clusters, and by infection Vertical lines are mean scores.

### Table S6. Patient management scores at baseline vs after intervention, by type of provider, by intervention/control clusters. SD = standard deviation

|  |  |  | **control** | | | | | | **intervention** | | | | | |
| --- | --- | --- | --- | --- | --- | --- | --- | --- | --- | --- | --- | --- | --- | --- |
|  |  |  | **baseline** | | | **endline** | | | **baseline** | | | **endline** | | |
| **site** | **Provider type** | **Infection scenario** | **visits** | **mean** | **SD** | **visits** | **mean** | **SD** | **visits** | **mean** | **SD** | **visits** | **mean** | **SD** |
| Both sites combined | community pharmacy | diarrhoea | 51 | 12·02 | 4·80 | 44 | 15·23 | 3·37 | 22 | 11·68 | 5·14 | 15 | 13·33 | 5·95 |
|  |  | fever | 32 | -0·69 | 1·82 | 35 | 0·71 | 2·76 | 11 | 0·55 | 2·46 | 13 | -0·31 | 1·55 |
|  |  | rhinitis/pharyngitis | 28 | 0·52 | 1·60 | 26 | 1·04 | 1·71 | 12 | 1·25 | 1·66 | 19 | 0·63 | 1·61 |
|  |  | severe pneumonia | 37 | 8·19 | 3·60 | 45 | 10·11 | 2·85 | 18 | 8·39 | 5·03 | 16 | 10·19 | 3·51 |
|  |  | uti | 20 | 6·10 | 2·99 | 27 | 7·93 | 2·6 | 7 | 3·43 | 2·76 | 13 | 6·69 | 4·27 |
|  | health centre | diarrhoea | 8 | 10·75 | 6·54 | 10 | 14·10 | 7·95 | 15 | 11·27 | 6·04 | 15 | 14·07 | 7·71 |
|  |  | fever | 13 | 6·77 | 3·90 | 10 | 6·20 | 4·18 | 18 | 5·22 | 3·56 | 13 | 6·46 | 3·71 |
|  |  | rhinitis/pharyngitis | 12 | 4·67 | 3·89 | 11 | 4·91 | 3·53 | 19 | 3·21 | 2·62 | 10 | 4·50 | 2·84 |
|  |  | severe pneumonia | 6 | 4·00 | 0 | 6 | 6·67 | 1·86 | 8 | 3·00 | 1·85 | 8 | 5·12 | 3·76 |
|  |  | uti | 11 | 8·27 | 4·94 | 13 | 7·77 | 4·53 | 18 | 8·17 | 4·83 | 17 | 8·24 | 5·39 |
|  | informal vendor | diarrhoea | 17 | -1·65 | 1·06 | 17 | -2·00 | 1·22 | 19 | -1·16 | 1·21 | 19 | -1·42 | 2·09 |
|  |  | fever | 17 | 0·29 | 0·85 | 17 | 1·82 | 0·53 | 19 | 0·21 | 0·71 | 19 | 1·68 | 0·48 |
|  |  | rhinitis/pharyngitis | 17 | 0·29 | 0·59 | 17 | 0·53 | 0·62 | 19 | 0·47 | 0·84 | 19 | 1·47 | 1·02 |
|  |  | severe pneumonia | 17 | 0·41 | 1·28 | 17 | 3·00 | 1·17 | 19 | 0·16 | 0·37 | 19 | 3·32 | 1·95 |
|  |  | uti | 17 | 1·12 | 0·78 | 17 | 1·00 | 0 | 19 | 0·89 | 0·88 | 19 | 0·84 | 0·69 |
| Kimpese | community pharmacy | diarrhoea | 50 | 12·16 | 4·74 | 43 | 15·47 | 3·02 | 19 | 13·05 | 3·64 | 12 | 15·83 | 3·24 |
|  |  | fever | 31 | -0·81 | 1·72 | 34 | 0·71 | 2·8 | 8 | -0·38 | 2·20 | 10 | -0·70 | 1·49 |
|  |  | rhinitis/pharyngitis | 27 | 0·54 | 1·63 | 25 | 1·04 | 1·74 | 9 | 1·22 | 1·56 | 16 | 0·38 | 1·50 |
|  |  | severe pneumonia | 36 | 8·31 | 3·58 | 44 | 10·27 | 2·67 | 15 | 9·67 | 4·43 | 13 | 11·46 | 2·30 |
|  |  | uti | 19 | 6·53 | 2·37 | 26 | 8·19 | 2·25 | 4 | 5·25 | 2·22 | 10 | 8·50 | 2·92 |
|  | health centre | diarrhoea | 2 | 20·00 | 5·66 | 4 | 23·00 | 2·16 | 7 | 16·29 | 5·09 | 7 | 21·57 | 3·15 |
|  |  | fever | 7 | 8·29 | 4·23 | 4 | 10·00 | 4·16 | 10 | 6·40 | 3·57 | 5 | 9·40 | 3·71 |
|  |  | rhinitis/pharyngitis | 6 | 6·83 | 3·87 | 4 | 8·75 | 2·36 | 11 | 4·45 | 2·07 | 3 | 7·67 | 2·52 |
|  |  | uti | 5 | 12·6 | 3·05 | 7 | 11·57 | 1·27 | 10 | 11·9 | 2·60 | 9 | 12·44 | 3·21 |
| Nanoro | community pharmacy | diarrhoea | 1 | 5·00 |  | 1 | 5·00 |  | 3 | 3·00 | 5·20 | 3 | 3·33 | 1·53 |
|  |  | fever | 1 | 3·00 |  | 1 | 1·00 |  | 3 | 3·00 | 1·00 | 3 | 1·00 | 1·00 |
|  |  | rhinitis/pharyngitis | 1 | 0·00 |  | 1 | 1·00 |  | 3 | 1·33 | 2·31 | 3 | 2·00 | 1·73 |
|  |  | severe pneumonia | 1 | 4·00 |  | 1 | 3·00 |  | 3 | 2·00 | 2·00 | 3 | 4·67 | 2·08 |
|  |  | uti | 1 | -200 |  | 1 | 1·00 |  | 3 | 1·00 | 0 | 3 | 0·67 | 0·58 |
|  | health centre | diarrhoea | 6 | 7·67 | 2·80 | 6 | 8·17 | 2·32 | 8 | 6·88 | 1·89 | 8 | 7·50 | 2·20 |
|  |  | fever | 6 | 5·00 | 2·83 | 6 | 3·67 | 1·37 | 8 | 3·75 | 3·15 | 8 | 4·62 | 2·39 |
|  |  | rhinitis/pharyngitis | 6 | 2·50 | 2·66 | 7 | 2·71 | 1·6 | 8 | 1·50 | 2·39 | 7 | 3·14 | 1·68 |
|  |  | severe pneumonia | 6 | 4·00 | 0 | 6 | 6·67 | 1·86 | 8 | 3·00 | 1·85 | 8 | 5·12 | 3·76 |
|  |  | uti | 6 | 4·67 | 2·66 | 6 | 3·33 | 1·86 | 8 | 3·50 | 1·77 | 8 | 3·50 | 2·51 |
|  | informal vendor | diarrhoea | 17 | -1·65 | 1·06 | 17 | -2 | 1·22 | 19 | -1·16 | 1·21 | 19 | -1·42 | 2·09 |
|  |  | fever | 17 | 0·29 | 0·85 | 17 | 1·82 | 0·53 | 19 | 0·21 | 0·71 | 19 | 1·68 | 0·48 |
|  |  | rhinitis/pharyngitis | 17 | 0·29 | 0·59 | 17 | 0·53 | 0·62 | 19 | 0·47 | 0·84 | 19 | 1·47 | 1·02 |
|  |  | severe pneumonia | 17 | 0·41 | 1·28 | 17 | 3 | 1·17 | 19 | 0·16 | 0·37 | 19 | 3·32 | 1·95 |
|  |  | uti | 17 | 1·12 | 0·78 | 17 | 1 | 0 | 19 | 0·89 | 0·88 | 19 | 0·84 | 0·69 |

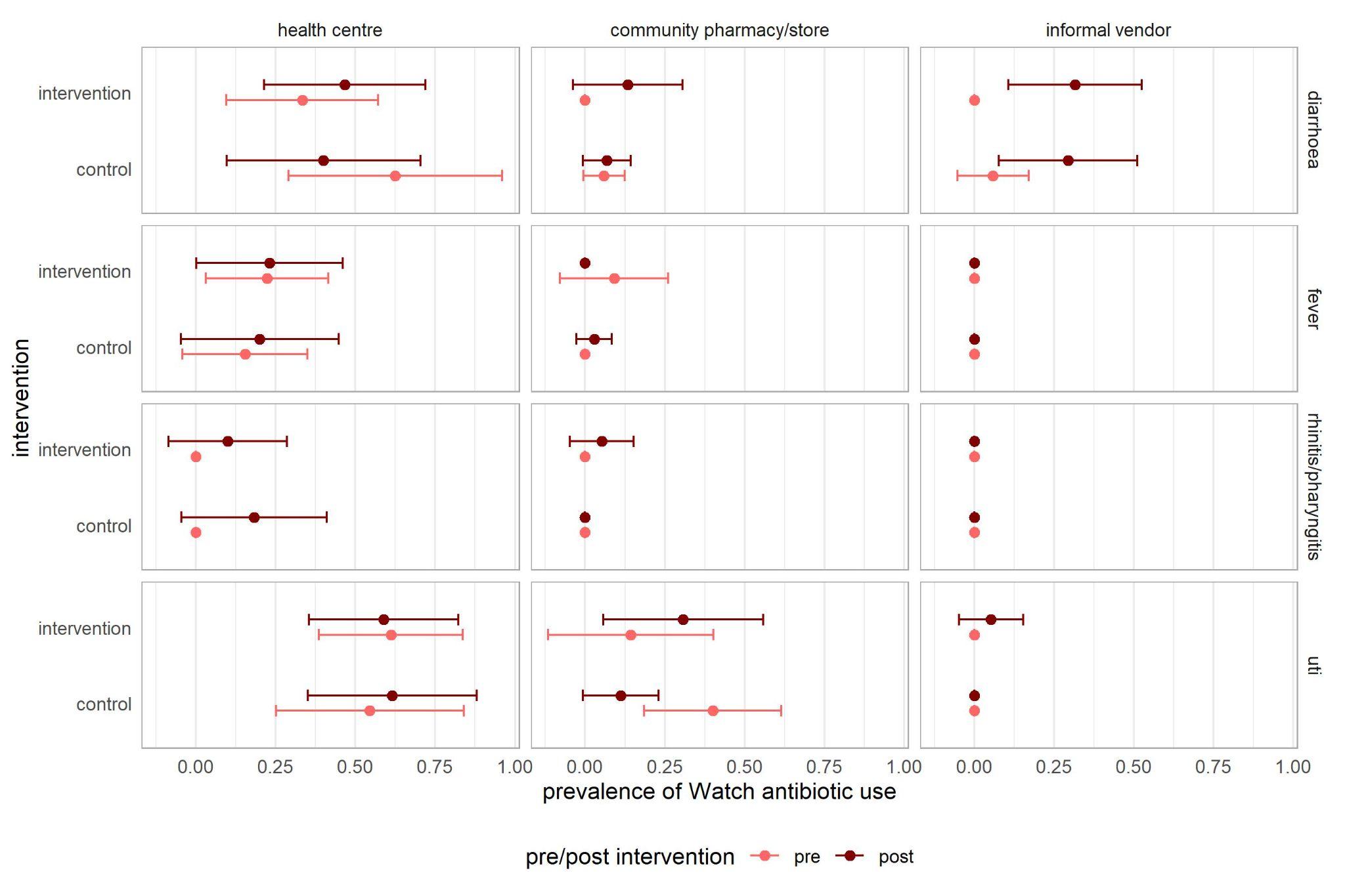

### Figure S5. Prevalence of Watch-group antibiotic use during simulated patient visits before and after the intervention, at providers in the intervention and control group

1. Ingelbeen B, Phanzu DM, Phoba M-F, Budiongo MYN, Berhe NM et al. (2022). Antibiotic use from formal and informal healthcare providers in the Democratic Republic of Congo: a population-based study in two health zones. *Clinical Microbiology and Infection*, *28*(9), 1272–1277. https://doi.org/10.1016/j.cmi.2022.04.002
2. Valia D, Kouanda JS, Ingelbeen B, Derra K, Kaboré B, et al. (2023). Healthcare seeking outside healthcare facilities and antibiotic dispensing patterns in rural Burkina Faso: A mixed methods study. *Tropical Medicine and International Health*, *28*(5), 391–400. https://doi.org/10.1111/tmi.13868
